# Systems Vaccinology Reveals Distinct Immune Signatures of Inhaled and Intramuscular SARS-CoV-2 Vaccination in Humans

**DOI:** 10.1101/2025.10.14.25337996

**Authors:** Lennart Riemann, Swantje Hammerschmidt, Rodrigo Gutierrez Jauregui, Ivan Odak, Joana Barros Martins, Ahmed Hassan, Leonie Marie Weskamm, Leonie Mayer, Verena Krähling, Rebekka Kraemer, Michaela Friedrichsen, Inga Ravens, Jasmin Ristenpart, Anja Schimrock, Gesine Hansen, Christine Falk, Stephan Becker, Asisa Volz, Gerd Sutter, Jens Hohlfeld, Marylyn M. Addo, Reinhold Förster

## Abstract

Mucosal vaccines may reduce both infection and transmission by engaging local immunity, yet the immunological pathways they activate in humans remain poorly defined. Here, we present a comprehensive systems vaccinology analysis of a Modified Vaccinia virus Ankara (MVA)-based SARS-CoV-2 vaccine candidate tested in two distinct phase 1 clinical trials as a booster vaccination with either inhaled or intramuscular delivery, and benchmarked findings against published mRNA vaccine data. Longitudinal multi-omics profiling of peripheral blood and bronchoalveolar lavage revealed that inhaled vaccination induces a distinct immunological signature, characterized by effector CD8+ T cell enrichment in the respiratory tract with minimal systemic perturbation. Inhaled delivery elicited limited systemic antibody responses, yet cellular immunity was comparable to that induced by intramuscular vaccination. Transcriptional profiling uncovered shared innate and proliferative programs across both delivery routes, with temporal decoupling of gene programs associated with humoral versus cellular immunity. Machine learning identified a robust early blood transcriptional signature predictive of antibody production. These findings offer a compelling molecular rationale for mucosal vaccine strategies and highlight actionable targets for optimizing mucosal vaccines. ClinicalTrials.gov identifier: NCT05226390, NCT04895449.

## Introduction

Most pandemics in the past century have been caused by respiratory pathogens that infect via mucosal surfaces.^1,2^ Yet nearly all licensed vaccines against them are administered intramuscularly, inducing robust systemic but limited mucosal immunity.^3–7^ This limits their ability to prevent infection and transmission—particularly against emerging variants—as demonstrated during the COVID-19 pandemic.^4,8,9^

Mucosal vaccines, including inhaled formulations, represent a compelling alternative. By mimicking natural infection, they activate local immune responses, potentially enabling early pathogen control, reduced transmission, and broader cross-reactivity.^3,10–12^ Additional advantages include dose-sparing potential, reduced systemic reactogenicity, and improved acceptability through needle-free delivery.^10,13–15^

Several inhaled vaccines have demonstrated protective efficacy in preclinical^11,16–19^ and clinical studies^20–24^. Notably, a head-to-head clinical trial showed that inhaled adenovirus-vectored Ad5-nCoV vaccination^21^—at one-fifth the intramuscular dose—elicited durable systemic and mucosal antibody responses, comparable T cell immunity, and a trend toward improved clinical protection against symptomatic COVID-19 with fewer adverse events.^22^ However, the systemic and local immune pathways engaged by inhaled vaccination—and how these differ from those elicited by intramuscular delivery—remain largely undefined in humans. Understanding these mechanisms is crucial for rational vaccine design and optimization.

Here, we applied a systems vaccinology approach to comprehensively profile immune responses elicited by an inhaled COVID-19 booster vaccination in humans. Using a Modified Vaccinia virus Ankara (MVA)-vectored COVID-19 vaccine candidate encoding a prefusion-stabilized Spike protein (MVA-SARS-CoV-2-ST; hereafter MVA-ST),^25^ we performed longitudinal multi-omics profiling at the cellular, proteomic, and transcriptomic levels in peripheral blood and bronchoalveolar lavage (BAL) fluid. MVA is a versatile and safe^26,27^ vaccine platform that is licensed^28,29^ or under clinical investigation^30–32^ for multiple pathogens, and therefore a clinically highly relevant model system to study. By comparing responses to inhaled versus intramuscular delivery of the same vaccine, and benchmarking against mRNA vaccine datasets, we reveal shared and distinct molecular signatures of mucosal and intramuscular vaccination, and identify an early gene expression signature predictive of antibody responses across platforms.

These findings provide a high-resolution map of mucosal vaccine-induced immunity and yield mechanistic insights to inform next-generation mucosal vaccine design.

## Results

### Study design, safety and immunogenicity of inhaled MVA-ST vaccination

We conducted a single-center, open-label phase 1 trial to assess safety, reactogenicity, and immunogenicity of the MVA-ST vaccine candidate, administered via inhalation at a dose of 10^7^ IU (Infectious Units) to 23 previously SARS-CoV-2 immunized adults (Fig. 1a). Detailed safety and immunogenicity outcomes have been reported elsewhere.^33,34^ Briefly, no serious vaccine-related adverse events occurred. One participant experienced a significant transient decline in pulmonary function, which resolved spontaneously without intervention. Humoral responses following inhaled MVA vaccination remained unchanged in plasma following vaccination, while cellular responses were robust, evidenced by increased IFN-γ production in whole blood following *ex vivo* SARS-CoV-2 Spike peptide stimulation (Fig. 1b).

**Figure 1:**
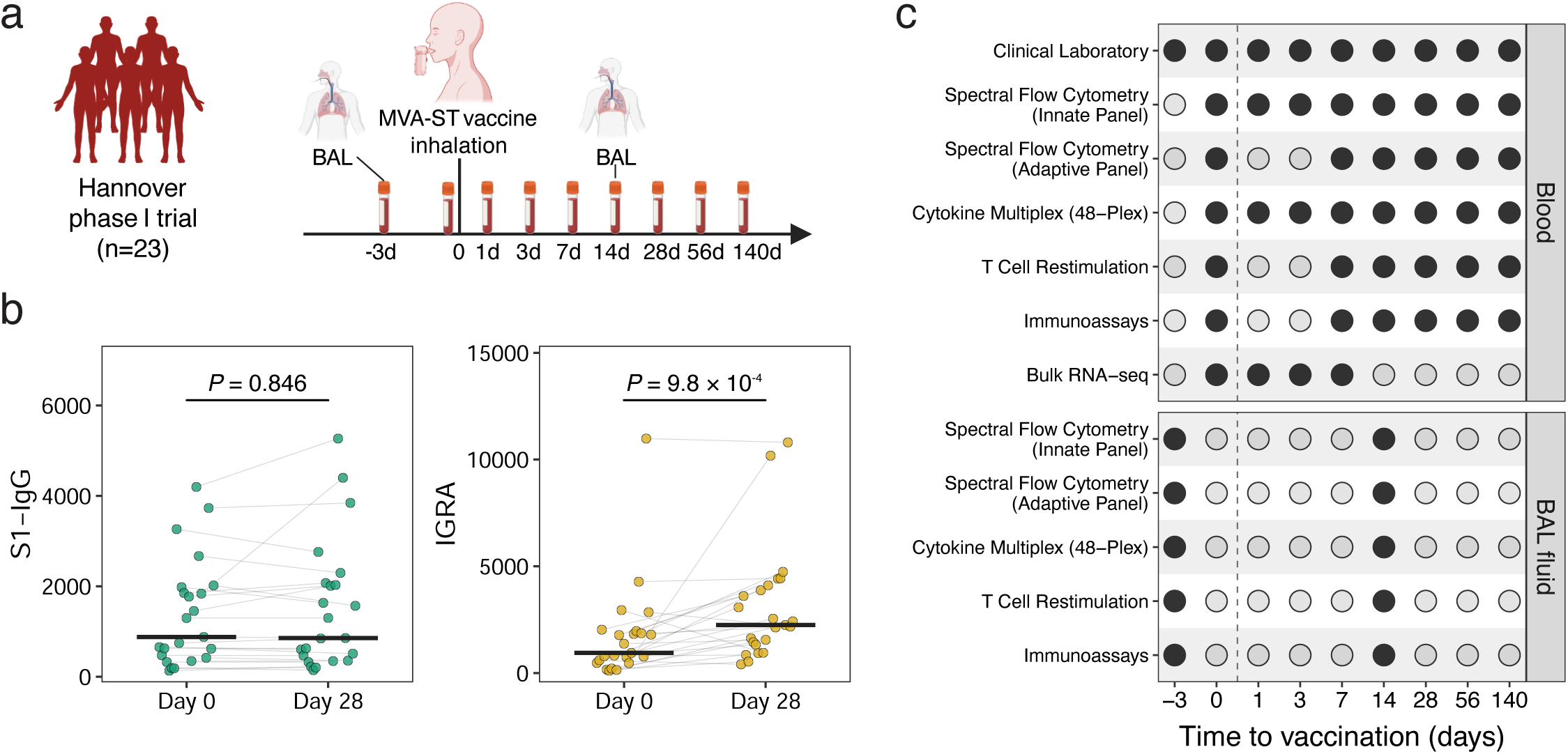
Study overview and vaccine responses to inhaled MVA-ST. **a**, Schematic overview of a systems-level immune profiling study of a Modified Vaccinia virus Ankara vaccine candidate encoding a prefusion-stabilized SARS-CoV-2 Spike protein (MVA-ST), administered via inhalation as part of a phase 1 clinical trial in healthy adults (Hannover cohort, n=23). Multi-omics profiling—including cellular, proteomic, and transcriptomic analyses—was conducted on blood and bronchoalveolar lavage (BAL) samples to characterize local and systemic responses. **b**, Systemic humoral and cellular immune responses before and after inhaled vaccination. Left, plasma S1-specific IgG concentrations at baseline and day 28. Right, IFN-γ release assay (IGRA) responses at baseline and day 28. Antibody levels remained unchanged, whereas IGRA responses increased significantly. Statistical comparisons were performed using paired Wilcoxon tests. **c**, Overview of biospecimen collection and multi-omics profiling performed in the inhaled vaccination cohort. Spectral flow cytometry, multiplex cytokine profiling and T cell restimulation assays were conducted on samples from both blood and BAL, whereas laboratory testing and bulk RNA sequencing were performed on blood samples only.

To comprehensively profile immune responses, we conducted a longitudinal systems vaccinology study collecting peripheral blood at baseline (day 0) and days 1, 3, 7, 14, 28, 56, and 140 post-vaccination, and BAL samples at days –3 (pre-vaccination) and +14 (Fig. 1c). We investigated cellular subsets, cytokine profiles, and antigen-specific T cell responses following *ex vivo* Spike peptide restimulation in both compartments, alongside whole-blood transcriptomic profiling at days 0, 1, 3, and 7 and monitoring of laboratory values.

Routine safety laboratory measurements remained within reference ranges throughout the 140-day study period.^33^ Complete blood counts showed no clinically meaningful deviations, with only a modest, transient rise in leukocyte and neutrophil counts on day 1 that remained within normal limits (Supplementary Fig. 1).

### Compartmentalized immune cell responses reveal selective CD8+ T cell enrichment in airways

High-dimensional spectral flow cytometry revealed distinct compartment-specific immune dynamics following inhaled vaccination. In BAL fluid, manual gating showed no significant changes in the proportions of phagocytic versus non-phagocytic cells, nor in the ratio of innate to lymphoid cells within the non-phagocytic fraction between baseline and day 14 (Supplementary Fig. 2a,b).

Unsupervised clustering of non-phagocytic cells using 27-parameter (innate) and 33-parameter (adaptive) panels identified 31 distinct clusters each, enabling deep immune phenotyping (Fig. 2a; Supplementary Fig. 2c; Supplementary Tables 1–2). While innate cluster frequencies exhibited no significant changes between baseline and day 14 (Supplementary Fig. 2d), one adaptive cluster (cluster 21) demonstrated significant enrichment at day 14, which persisted after correction for multiple testing (Fig. 2b,c).

**Figure 2:**
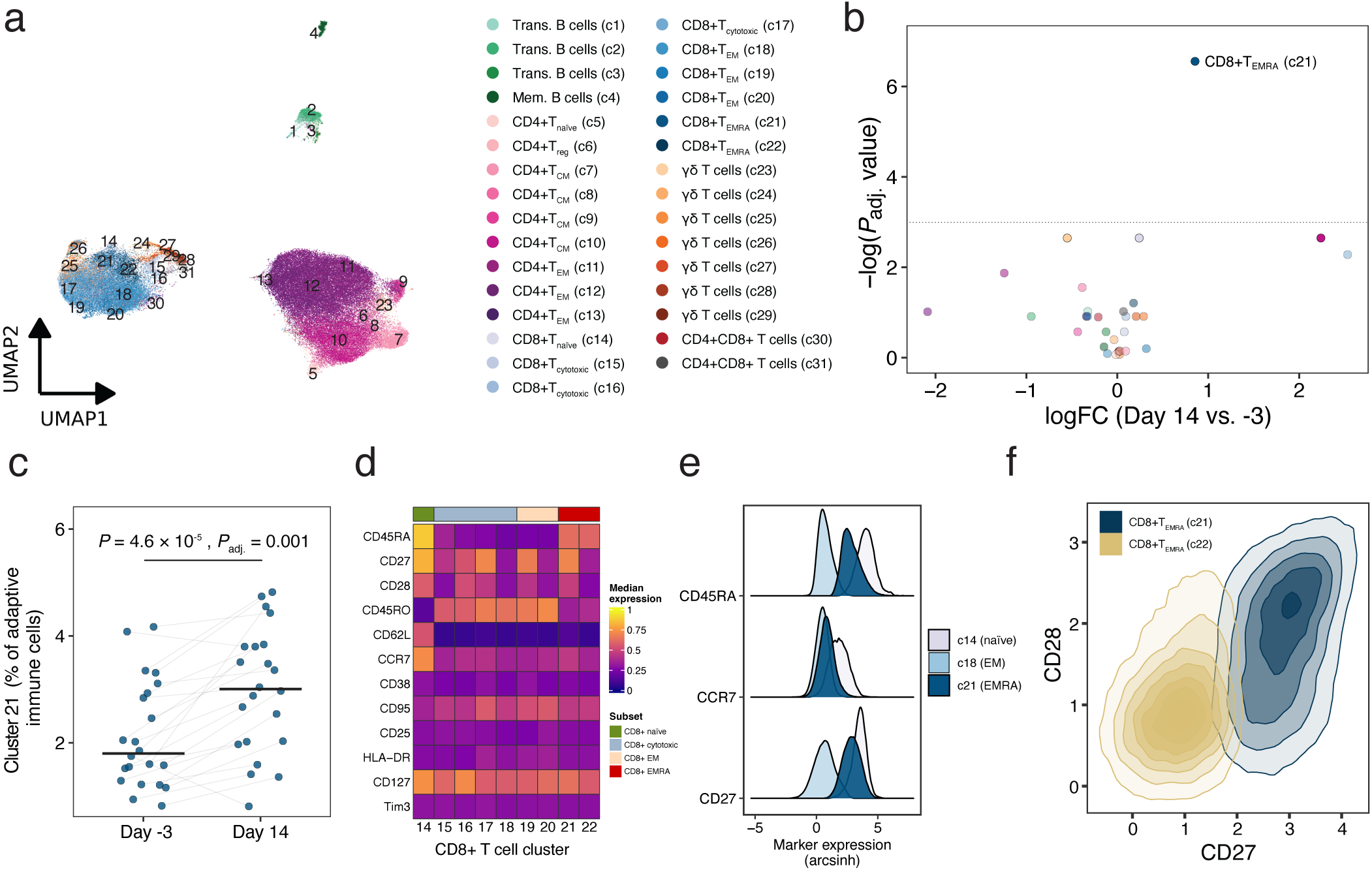
Immune cell changes in bronchoalveolar lavage fluid after inhaled vaccination. **a**, UMAP projection of 31 adaptive immune clusters in non-phagocytic BAL cells. **b**, Volcano plot of adaptive cluster abundance changes between day 14 and day –3. **c**, Frequencies of significantly enriched cluster 21 as a proportion of adaptive BAL cells at day –3 and day 14. *P* value derived from the differential abundance analysis. **d**, Heatmap showing the expression of canonical T cell markers in the CD8 T cell clusters (clusters 14–22). **f,** Ridge plot showing expression of CD45RA, CD27, and CCR7 in cluster 21 (blue) compared to naïve and EM CD8+ T cell clusters in BAL (grey), indicating a T_EMRA_-like phenotype with preserved co-stimulatory molecule expression. **e**, 2D cytometry plot comparing CD27 and CD28 expression in cluster 21 versus phenotypically similar cluster 22, highlighting characteristic expression of co-stimulatory molecules in cluster 21.

This enriched population displayed a CD8+CD45RA+CCR7-CD27+CD28+ phenotype, consistent with T_EMRA_-like cells (effector memory T cells re-expressing CD45RA) that retained co-stimulatory molecules CD27 and CD28—markers associated with preserved effector and reactivation potential (Fig. 2d,e; Supplementary Table 2). Notably, a phenotypically similar cluster (cluster 22) lacking CD27 and CD28 showed no frequency changes, indicating selective enrichment of the co-stimulatory molecule-positive subset (Fig. 2f). These findings suggest targeted expansion or recruitment of antigen-experienced CD8+ T cells with less terminal differentiation following mucosal vaccination.

In contrast, immune profiling of peripheral blood using identical antibody panels revealed minimal changes in systemic immune cell composition. Among 31 innate and 30 adaptive clusters, only one small neutrophil subset transiently reached statistical significance at day 7 (Supplementary Fig. 3a–d; Supplementary Tables 3–4). Notably, no other innate or adaptive immune cell clusters, including B and T cell subsets, displayed significant frequency changes systemically at any time point.

These findings demonstrate that inhaled vaccination induces localized, targeted enrichment of phenotypically distinct T_EMRA_-like CD8+ T cells in the respiratory tract without perturbing systemic immune cell frequencies, consistent with compartmentalized mucosal immune responses.

### Early peripheral chemokine suppression contrasts with robust local recall responses

Multiplex analysis of 48 plasma cytokines revealed an early and transient decrease in multiple chemokines and growth factors following inhaled vaccination (Fig. 3a). Specifically, CCL5, CCL11, CCL27, CXCL12, MIF, SCGF-β and PDGF-BB decreased significantly on days 1–3 relative to baseline, subsequently returning to baseline by day 7 (Fig. 3b, Supplementary Fig. 4a). IL-7 levels rose slightly in some participants on day 1–3, but no consistent pattern emerged across the cohort. Importantly, canonical pro-inflammatory cytokines—such as TNF-α, IFN-γ, CXCL9, and CXCL10—remained unchanged, contrasting with the cytokine surges reported after multiple intramuscular vaccines, including mRNA vaccines.^35–40^

**Figure 3:**
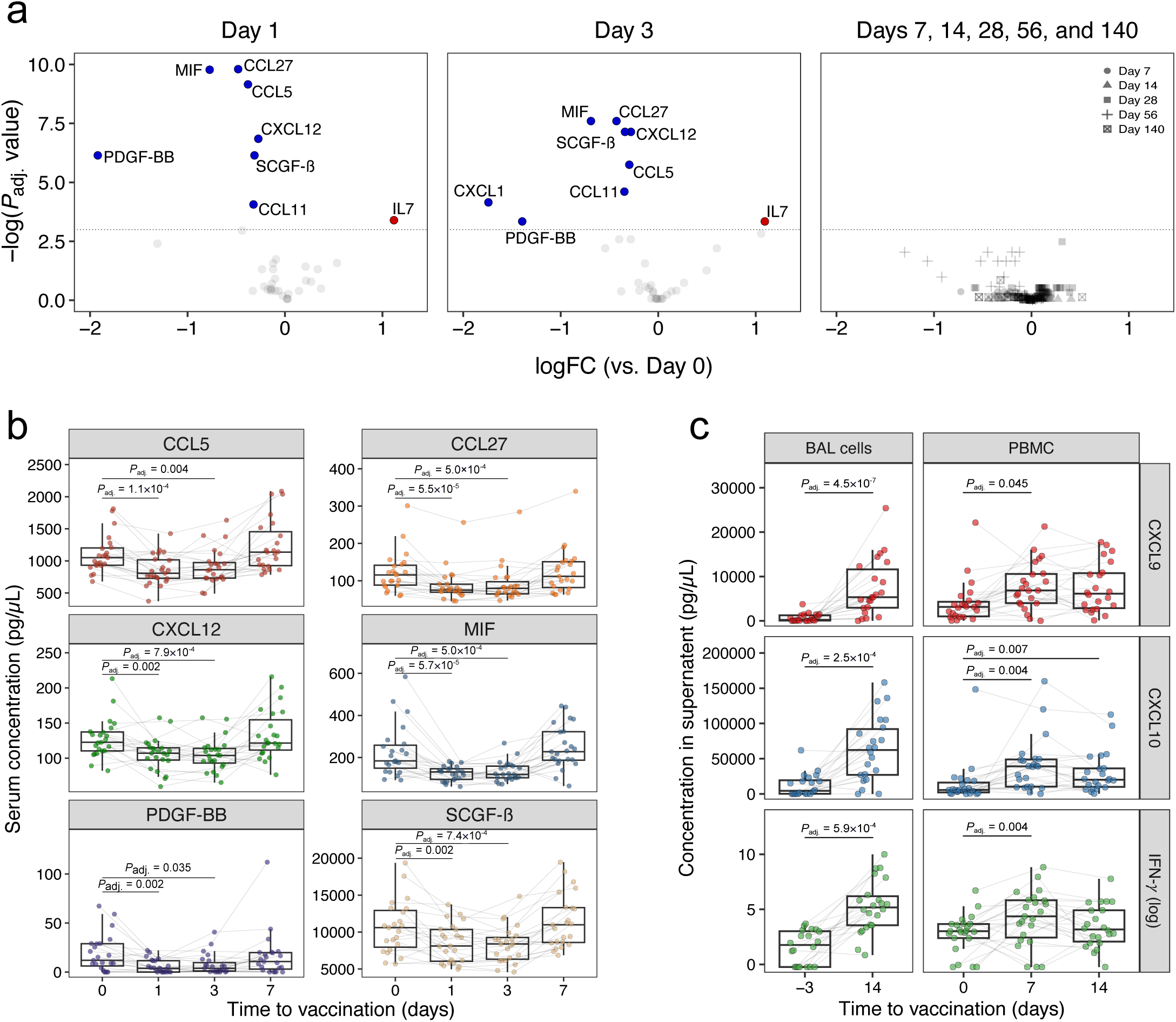
Distinct cytokine dynamics in plasma and antigen-stimulated cultures following inhaled vaccination. **a**, Volcano plots depicting differential cytokine abundance in plasma at day 1 (left), day 3 (middle), and at later time points (days 7–140, right) relative to baseline. Analytes with significant increases (adjusted *P* <0.05) are shown in red, and those with significant decreases in blue. Dashed lines indicate significance thresholds. **b**, Early concentration profiles of selected plasma cytokines that were significantly altered at days 1 and 3 compared to baseline, showing a transient decrease that returned to baseline by day 7 and remained stable thereafter (Supplementary Fig. 4b). **c**, Concentrations of CXCL9, CXCL10, and IFN-γ in culture supernatants from BAL cells and PBMCs after *ex vivo* restimulation with a SARS-CoV-2 Spike peptide pool. Only the early time points are shown; later time points (only available for PBMCs) are presented in Supplementary Fig. 4c. The change from baseline to day 14 was substantially greater in BAL cells than in PBMCs. Statistical testing in all panels was performed using linear modeling with limma; multiple testing correction was applied independently at each time point.

To investigate whether these plasma protein changes reflected altered gene expression, we analyzed mRNA levels of the significantly altered chemokines and growth factors in whole blood. Despite the observed plasma protein decreases, corresponding transcript levels showed no significant changes at any time point (Supplementary Fig. 4b), suggesting that reduced plasma concentrations primarily reflect redistribution or consumption of circulating chemokines rather than transcriptional suppression at this dose level.

In BAL fluid, most cytokines remained below detection thresholds, with only 10 analytes detectable in >50% of samples. CXCL10 levels increased slightly at day 14 (*P* = 0.022), but this change did not retain statistical significance after multiplicity correction (*P*_adj._ = 0.597; Supplementary Table 5).

Functional assessment through *ex vivo* Spike peptide restimulation revealed a striking contrast between local and systemic recall responses. BAL cells collected at day 14 secreted significantly higher levels of CXCL9, CXCL10, and IFN-γ levels compared to baseline, confirming potent local memory T cell responses within the lung (Fig. 3c). In contrast, peripheral blood mononuclear cells (PBMCs) mounted weaker responses that peaked at day 7 and waned thereafter (Fig. 3c, Supplementary Fig. 4c). The magnitude of the BAL response exceeded that of PBMCs, underscoring the predominance of mucosal over systemic T cell immunity.

Collectively, these findings reveal that inhaled vaccination selectively modulates peripheral chemokines and growth factors without triggering systemic inflammatory cytokine responses, while enhancing local antigen-specific T cell memory responses in the lung.

### A transient innate transcriptional program precedes adaptive activation

Bulk RNA-sequencing of whole-blood samples revealed a dynamic transcriptional response to inhaled vaccination with distinct temporal phases. At day 1, 911 genes were upregulated and 599 downregulated (false discovery rate, FDR <0.05), followed by a quiescent phase at day 3 and a smaller transcriptional wave at day 7 (136 upregulated, 67 downregulated genes; Fig. 4a).

**Figure 4:**
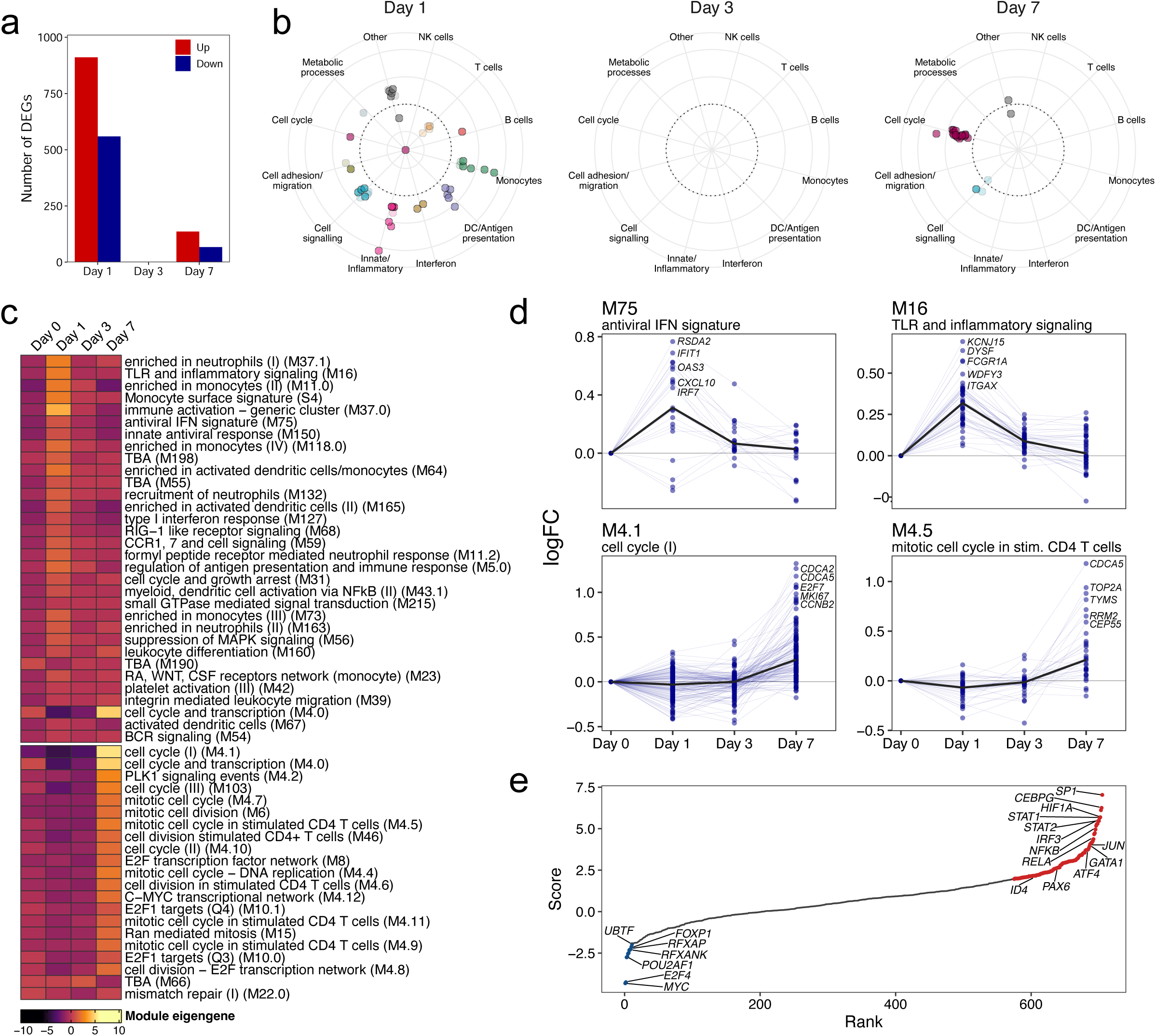
Transcriptional profiling reveals temporal immune dynamics and lung T cell correlates following inhaled vaccination. **a**, Number of differentially expressed genes (FDR <0.05) at days 1, 3, and 7, relative to baseline. **b**, Polar plots depicting module eigengene changes of blood transcription modules (BTMs) at days 1, 3, and 7 versus baseline. Each dot represents a module; module eigengenes are defined as the first principal component of gene expression within each module and represent the overall module activity. Radial distance from the center indicates the magnitude of change relative to baseline: modules outside the dashed circle are upregulated; and those inside are downregulated. Dot opacity indicates significance (FDR <0.05: opaque; <0.1: semi-transparent). Modules are arranged angularly for visual separation; the plot highlights which immune programs are significantly modulated at each time point. Only significantly enriched modules are shown **c**, Heatmap of BTM eigengene expression at days 0, 1, 3, and 7 for significantly enriched modules. **d**, Median gene-level fold change trajectories over time for four selected modules representative of key immune programs. **e**, Transcription factor enrichment analysis showing ranked scores for inferred regulator activity at day 1. Red and blue indicate significantly increased or decreased transcription factor activity, respectively (*P* <0.05).

Mapping these signatures onto blood transcription modules (BTMs)^41^ revealed early systemic innate activation, with interferon, monocyte, and inflammatory modules peaking at 24h (Fig. 4b–d). By day 3, these innate signatures had returned to baseline, indicating a rapid but transient innate activation. At day 7, proliferative and adaptive modules emerged that were largely absent at earlier time points, demonstrating a coordinated transition from innate to adaptive activation.

Consistent with the BTM analysis, transcription factor enrichment analysis at day 1 (vs. day 0) highlighted increased activity of *STAT1*, *STAT2*, *NF-*κ*B* family members and *SP1*, reflecting inflammatory signaling and antigen-presenting cell priming (Fig. 4e). Conversely, transcription factors governing cell cycle regulation (*E2F4*, *MYC*) and homeostasis (*FOXP1*) were repressed, delineating a coordinated innate transcriptional program that engages pro-inflammatory pathways while temporarily suppressing proliferative pathways.

To explore functional connections between early systemic transcriptional responses and lung-localized T cell immunity, we correlated gene module changes with frequencies of Spike-specific IFN-γ-producing CD4+ and CD8+ T cells in BAL fluid collected at day 14 post-vaccination. At day 1, modules related to cell migration and adhesion correlated positively with both T cell subsets, while mitosis-related and specific chemokine modules showed inverse associations (Supplementary Fig. 5). On day 3, type 1 interferon modules remained significantly associated with both T cell responses. More correlations were observed for CD8+ T cell responses compared to CD4+ T cells across the early time points. By day 7, proliferative modules—particularly those linked to CD4+ T cells—showed the strongest positive associations, suggesting that early innate cues prime lung-resident T cell responses that are subsequently amplified by a proliferative wave one-week post-vaccination.

### Comparative transcriptional dynamics following inhaled versus intramuscular MVA-ST vaccination

To evaluate systemic immune signatures elicited by inhaled versus intramuscular booster vaccination, we compared blood transcriptomes from the inhaled MVA cohort to those of a parallel phase 1b trial evaluating intramuscular MVA-ST administration at escalating doses (low, medium, high IM groups; low dose equivalent to the inhaled dose; Fig. 5a). Analysis of immunological outcomes revealed that S1-specific IgG log_2_-fold changes were significantly lower following inhaled vaccination compared to all three IM dose levels, with no significant differences observed within IM groups (Supplementary Fig. 6a). In contrast, IFN-γ release assay log_2_-fold changes notably showed no significant differences across any of the four vaccination regimens, indicating that cellular immune responses were comparably induced regardless of delivery route or dose (Supplementary Fig. 6b). Both trials employed harmonized protocols, enabling direct comparisons across routes and doses following batch-correction (Supplementary Fig. 6c,d).

**Figure 5:**
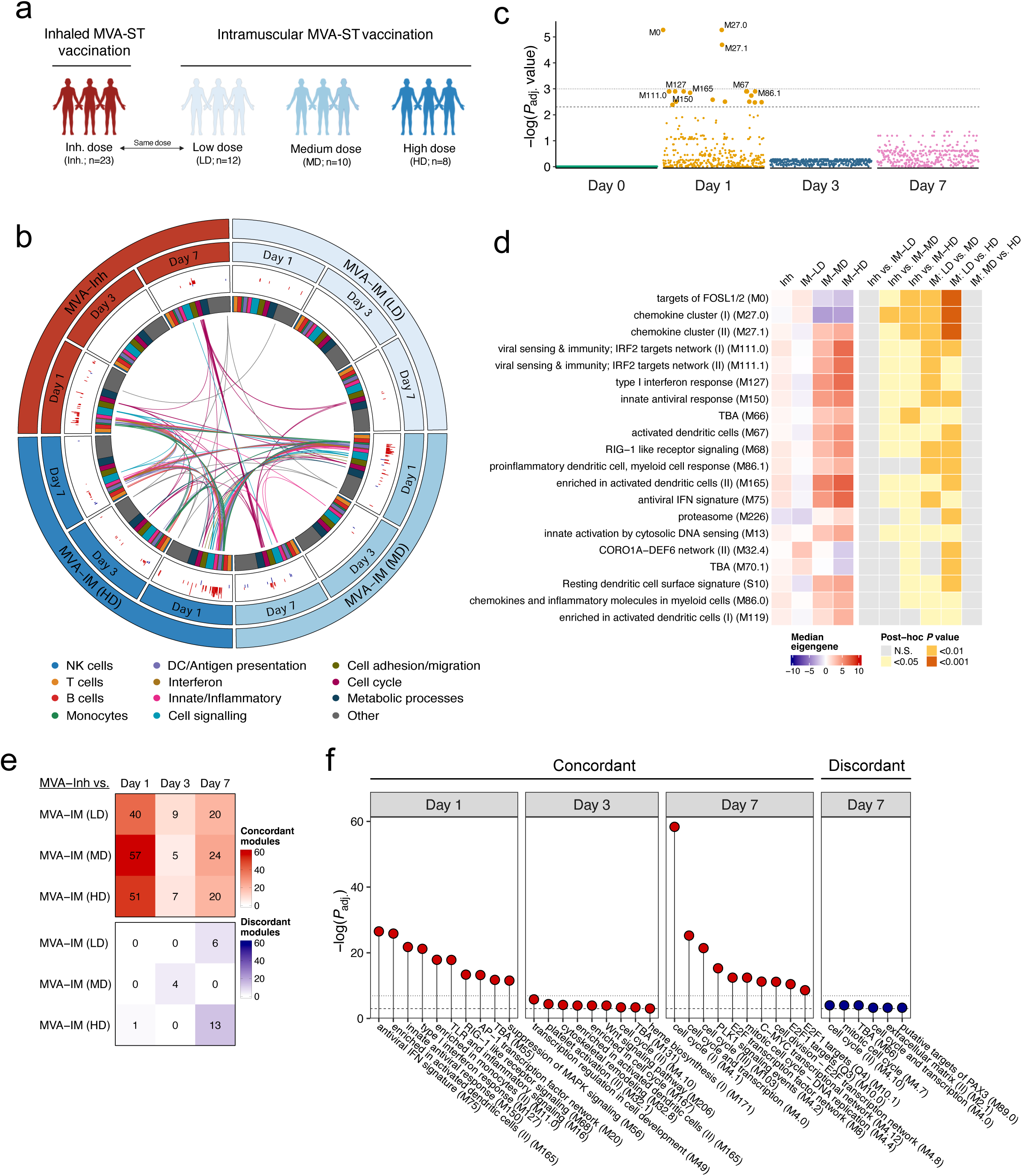
Comparative transcriptional profiling of inhaled versus intramuscular MVA-ST vaccination. **a**, Schematic overview of the inhaled vaccine group and the three IM vaccine groups included in the comparison. Immunological outcomes for these groups are shown in Supplementary Fig. 6a,b. **b**, Circos plot showing overlap of significantly enriched BTMs between inhaled and IM groups (low, medium, high dose) at days 1, 3, and 7. Each segment represents one group; lines connect BTMs with shared enrichment. Line colors and inner tracks denote functional BTM categories. Bar heights reflect the magnitude and direction of module expression changes (red: upregulated, blue: downregulated vs. baseline). **c**, Scatter plot showing module-level differential expression across vaccine regimens at days 0, 1, 3, and 7, based on Kruskal-Wallis tests of blood transcriptional module eigengene expression. Each dot represents one transcriptional module; dotted and dashed lines indicate FDR thresholds of 0.1 and 0.05, respectively. *P* value adjustment was performed independently for each time point. **d**, Heatmap of module eigengene expression (left) and adjusted *P* values from Dunn’s post-hoc tests (right) for BTMs that differed significantly across vaccine groups at day 1, as identified in Fig. 5c. **e**, Differential concordance analysis quantifying the number of modules that were concordantly or discordantly regulated between inhaled and intramuscular vaccination groups at days 1, 3, and 7. **f**, Top 10 (or fewer if not significant) concordant and discordant BTMs between inhaled and dose-matched low-dose IM groups across days 1, 3, and 7. Only day 7 showed significant discordant modules and is therefore displayed.

At day 1, inhaled and low-dose IM groups shared minimal overlap in enriched BTMs, with only a single co-enriched module (leukocyte differentiation), whereas broader overlap emerged between inhaled and medium/high IM groups that largely reflected innate sensing and antigen-presentation pathways (Fig. 5b). At day 7, shared enrichment of cell-cycle modules emerged, particularly between the inhaled and medium-dose IM cohorts. Notably, the highest IM dose maintained several inflammatory modules from day 1 that had resolved in other groups.

Quantitative analysis of module expressions using eigengene values (representing overall module activity^42^) across all 346 modules showed no significant differences between cohorts at baseline, days 3, or day 7 (Fig. 5c). At day 1, however, 21 modules differed across groups (FDR <0.1), with 3 modules reaching FDR <0.05—all related to innate immune activation.

Pairwise comparisons showed no significant differences between inhaled and low-dose IM groups, but revealed a clear dose-response gradient across the IM groups, with the highest expression observed in the high-dose group (Fig. 5d). Interestingly, one chemokine cluster showed significantly lower expression in the medium– and high-dose IM groups than in the inhaled and low-dose IM groups.

We next applied differential concordance analysis to explicitly delineate pathways with coherent or opposing kinetics across regimens. On day 1, 40–51 modules were concordantly regulated between the inhaled and respective IM groups, dominated by innate immune programs (Fig. 5e,f). Concordance diminished by day 3, then re-emerged around proliferative modules at day 7. Discordant modules were rare and largely limited to day 7 comparisons, and the statistical associations were substantially weaker. Some of these modules appeared in both concordant and discordant lists, indicating divergent kinetics within a module.

These results demonstrate that inhaled and intramuscular vaccination engage similar early innate and later proliferative transcriptional programs, modulated by antigen dose and with only subtle route-specific differences.

### Distinct transcriptional correlates underpin antibody and T cell responses

To identify transcriptional predictors of adaptive immunity, we leveraged harmonized measurements of S1-specific IgG titers and T cell IFN-γ release at baseline and day 28 across both trials, correlating longitudinal BTM changes with immunological outcomes.

Overall, correlations were modest—consistent with the multifactorial nature of vaccine responsiveness—but distinct temporal and immunological patterns emerged (Supplementary Fig. 7a,b). Antibody titers showed strongest associations with transcriptional signatures on day 1, with 48 significantly correlated modules, primarily linked to innate and antiviral pathways. These early antibody-associated modules notably did not overlap with those linked to T cell responses. By day 3, antibody-related correlations decreased in both number and effect size, with only 7 modules correlating at day 7.

In contrast, IFN-γ release showed minimal module associations at day 1, but peaked at day 3 with 30 significantly correlated modules. Several exhibited negative associations and were linked to metabolic and transcriptional regulation pathways, suggesting that transient suppression of homeostatic programs supports effective T cell priming. Of note, very few modules were jointly associated with both humoral and cellular responses at any time point.

We further investigated whether pre-vaccination transcriptional states could predict subsequent immunogenicity (Supplementary Fig. 7c). Twenty-three baseline BTMs showed significant correlations, with most inversely associated with IFN-γ release, while baseline features were largely uninformative for antibody responses.

This integrative analysis reveals temporal uncoupling between transcriptional programs governing humoral versus cellular immunity, with early innate module induction correlating with antibody response and later transcriptional shifts plus pre-existing states correlating with T cell responses.

### Cross-platform benchmarking reveals platform-specific immune signatures

To contextualize inhaled MVA-ST immune responses, we benchmarked transcriptional profiles against COVID-19 mRNA vaccines using publicly available datasets spanning primary, secondary, and tertiary doses (Supplementary Table 6). This comparison, following data integration (Supplementary Fig. 8a,b), aimed to understand platform-specific immune activation patterns and their relationship to antibody responses.

Comparative analysis revealed that second and third mRNA booster doses—but not primary vaccination—elicited markedly stronger activation of innate immune modules than inhaled MVA (Fig. 6a). The median log_2_-fold change in type I interferon and viral sensing modules was over two-fold higher in mRNA booster recipients compared to inhaled booster MVA or primary mRNA groups (Fig. 6b).

**Figure 6:**
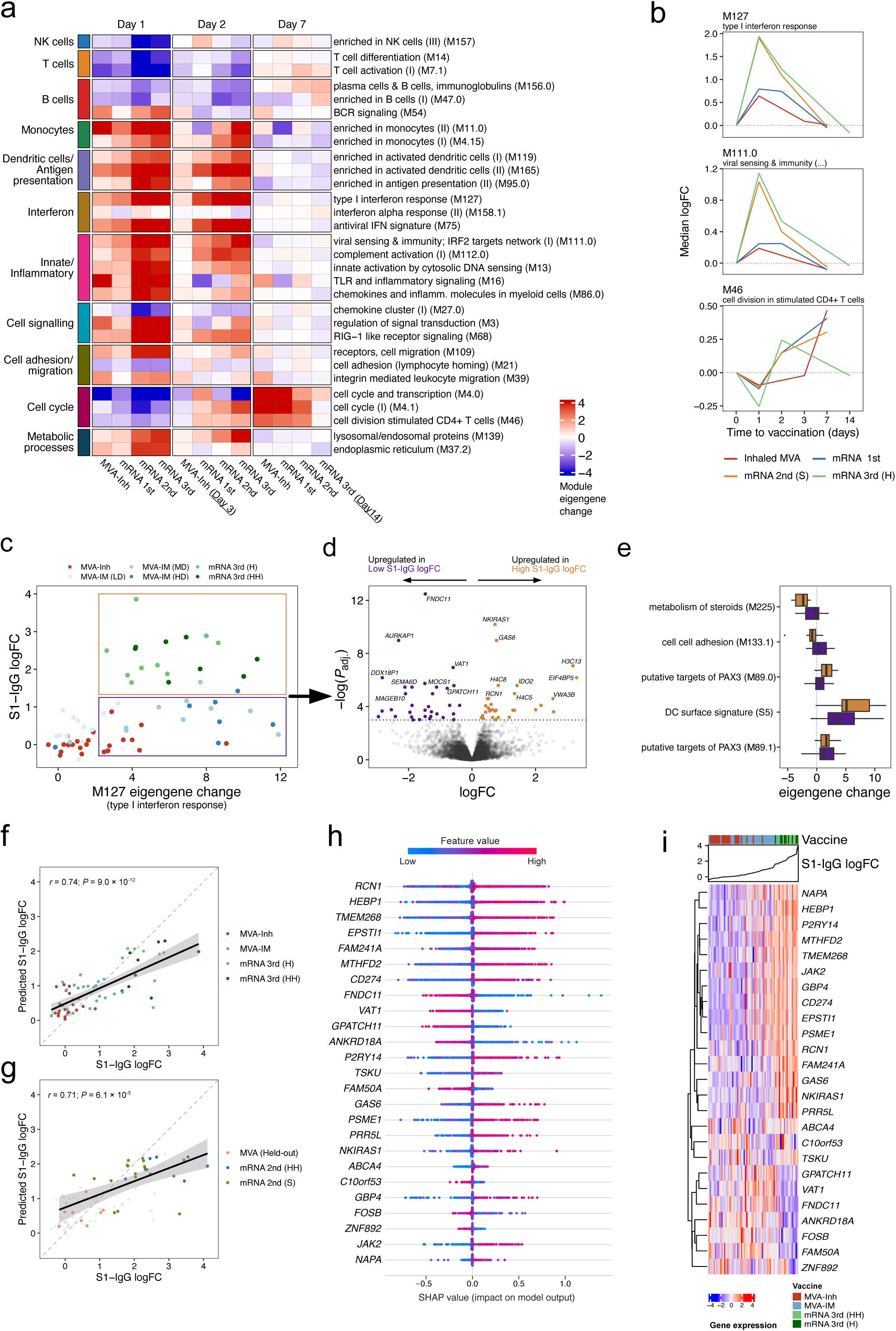
Cross-platform integration and predictive modeling of antibody responses. **a**, Heatmap showing longitudinal changes in BTM eigengene expression relative to baseline (day 0) in inhaled MVA and mRNA vaccine cohorts (first, second, and third doses). **b**, Median log_2_-fold change over time for three representative modules, highlighting shared and distinct dynamics across cohorts. **c**, Relationship between day 1 induction of a type I interferon response module (M127) and S1-specific antibody response at day 28. Among participants with robust IFN responses, both high (yellow box) and low (purple box) antibody responders were identified for subgroup analysis. **d**, Differential gene expression analysis comparing transcriptional changes from day 0 to day 1 between high (yellow) and low (purple) antibody responders within the robust IFN-response subgroup. The volcano plot shows genes with significant differences in day 1 induction between groups. The dashed line indicates FDR < 0.05. **e,** Blood transcriptional module enrichment analysis of differentially expressed genes. Only five modules were significantly enriched, though differences were modest, suggesting further gene-level investigations may be more informative. **f**, Machine-learning-based prediction of S1-specific IgG responses. Scatter plots compare observed versus predicted log_2_-fold changes in S1-specific IgG titers (day 28 versus day 0), based on average out-of-fold (OOF) predictions from iterative cross-validation on the training dataset. Each dot represents one individual; color denotes cohort. **g**, Predictions on an independent test set comprising 8 held-out MVA recipients and external mRNA vaccine cohorts (second dose). Second-dose mRNA data were used for independent validation as third-dose datasets were incorporated into the training set. **h**, Summary of the top 25 most influential gene features from the final XGBoost model, ranked by SHAP values. Each dot represents a participant (averaged across iterative cross-validation folds), with color indicating the feature value and position on the x-axis denoting its impact on the model output. **i**, Heatmap of z-score normalized expression of the top 25 SHAP-identified genes across all participants. Columns are ranked by observed S1-specific IgG log_2_-fold change (left to right), and rows (genes) are hierarchically clustered. Figure 6i validates the biological relevance of machine-learning derived features by demonstrating coherent expression patterns that correlate with antibody outcomes across the full participant spectrum.

Across all groups, innate module induction peaked on day 1, and declined by days 2–3, returning to baseline by day 7 (Fig. 6a). Modules associated with NK/T cell function and chemokine signaling were transiently suppressed in all groups but showed more pronounced downregulation in the mRNA booster cohorts. A CD4+ T cell-associated cell cycle module notably dipped at day 1 before rebounding at day 7 across platforms (Fig. 6b).

Extending correlation analysis between day 1 transcriptional activity and antibody responses (Supplementary Fig. 7) to mRNA cohorts, we found that 9 of 10 top-performing modules from the previous MVA-only analysis retained significant, directionally consistent correlations in the combined dataset (Supplementary Fig. 8c). Strikingly, several innate immunity modules displayed a heteroscedastic relationship: individuals with weak module induction (e.g., type I interferon response, M127) consistently mounted poor antibody responses, while those with strong induction showed a wide range of outcomes (Fig. 6c). This pattern suggests robust innate activation is necessary but not sufficient for strong humoral immunity.

Comparing the difference in gene expression between baseline and day 1 in high versus low antibody responders among individuals with robust innate induction identified 32 genes upregulated in high responders and 33 enriched in low responders, with modest module enrichment signals including dendritic cell surface genes in high responders and steroid metabolism in low responders (Fig. 6d,e). While these module-level analyses support detectable transcriptional differences between response groups, both the dichotomization of individuals into categories and the aggregation of genes into modules may mask more nuanced, gene-level associations across the full, continuous spectrum of antibody responses.

### Machine learning unveils gene-level correlates of vaccine-induced antibody responses

To capture finer-scale, gene-level predictors more precisely, we next employed a machine learning approach leveraging day 1 log_2_-fold changes from 1000 curated genes to model antibody responses as a continuous outcome (Supplementary Fig. 9a; see also Methods). After batch correction (Supplementary Fig. 9b,c), we trained an Extreme Gradient Boosting (XGBoost)^43^ model on 61 previously vaccinated individuals across the MVA (inhaled and IM) and two mRNA cohorts following booster administration.

The model demonstrated robust cross-validated predictive performance, indicating strong correlation between predicted and actual antibody responses (Pearson’s r = 0.74, 95% CI 0.58–0.85; *P* = 9.0×10^-12^; RMSE = 0.71; MAE = 0.54; Fig. 6f). Importantly, the model retained predictive ability on independent validation datasets comprising 8 held-out MVA vaccine recipients and 26 individuals from two distinct mRNA second-dose cohorts (Pearson’s r = 0.63, 95% CI 0.36–0.83; *P* = 6.1×10^-5^; RMSE = 0.94; MAE = 0.69; Fig. 6g).

While the model showed a tendency to overestimate responses in low responders and underestimate high responders, the consistent performance across both internal and external validation demonstrates its capacity to capture stable gene-level patterns quantitatively linked to downstream antibody responses.

To mechanistically interpret the driving predictive features, we analyzed SHAP (SHapley Additive exPlanations)^44^ values, which quantify how much each gene contributes to individual antibody response predictions. Top-ranked genes included known immune regulators and novel candidates (Fig. 6h; full list in Supplementary Material 1). Examination of their individual-level expression patterns further confirmed the biological relevance of the machine learning model, showing that participants with stronger antibody responses exhibited the expected coordinated upregulation of these key predictive features (Fig. 6i). Analysis of SHAP values for representative individual participants further illustrated how specific gene expression patterns contribute to the predicted antibody responses across cohorts (Supplementary Fig. 9d-f).

Beyond interferon-inducible genes (*JAK2*, *EPSTI1* and *GBP4*), positively associated genes included several regulators of immune balance such as RCN1, a multifunctional protein involved in stress responses,^45^ *MTHFD2*, a key metabolic checkpoint regulating T cell activity,^46^ and *CD274* (PD-L1), an immune checkpoint molecule suppressing T cells.^47^ *HEBP1*, which generates chemoattractant peptides for myeloid cells,^48^ also emerged as a key predictor. Negatively associated genes, such as *FNDC11*, *VAT1*, and *GPATCH11,* may reflect suppressive programs limiting humoral responses. Multiple poorly characterized genes (*TMEM268*, *ABCA4* and *FAM241A*) displayed high SHAP importance, nominating them for mechanistic investigation. Several top SHAP-ranked genes overlapped with the differentially expressed genes from the previous analysis (Supplementary Fig. 8e), providing orthogonal support for their relevance in shaping antibody responses.

These findings show that predictive modeling based on early blood transcriptomes can uncover robust and interpretable gene signatures of vaccine-induced antibody magnitude. Beyond established innate defense pathways, the analysis highlights broader regulatory architecture encompassing metabolic, checkpoint, and chemotactic genes that governs effective humoral immunity and offers mechanistic targets for vaccine optimization.

## Discussion

Systems vaccinology has illuminated the molecular underpinnings of vaccine immunogenicity across various platforms and pathogens such as yellow fever^49^, influenza^50–52^, and SARS-CoV-2^39,40,53^, yet nearly all studies have focused on parenteral vaccine delivery. Here, we present the first systems-level analysis of a mucosal SARS-CoV-2 vaccine in humans, profiling cellular, cytokine, and transcriptional responses after inhaled MVA-based vaccination.

Inhaled MVA-ST vaccination elicited a distinct immunological signature, with limited antibody responses but robust local and systemic T cell immunity, including selective enrichment of CD8+ T_EMRA_ cells in the lung. This compartmentalized pattern aligns with preclinical models of inhaled MVA.^17^ Peripheral pro-inflammatory cytokine and immune cell changes were minimal, yet antigen-specific T cell recall responses were stronger in BAL cells than in PBMCs. This profile—potent T cell immunity with attenuated systemic inflammation—supports mucosal vaccination strategies.

Transcriptional kinetics closely paralleled those observed across 13 parenteral vaccines,^54^ indicating conserved temporal patterns across pathogen, platform, and delivery route. Gene module responses were largely concordant with intramuscular delivery of the same vaccine. Although inhaled vaccination induced more modules at matching doses—possibly reflecting the lung’s extensive surface area and vascularization^55^—systemic antibody responses remained lower than all intramuscular doses, while cellular immunity was comparable across regimens. Whether higher inhaled doses could enhance humoral responses warrants further study.

Previous studies focused primarily on transcriptional predictors of humoral responses.^41,54,56,57^ Our study extends this work by analyzing humoral and cellular immunity in parallel, correlating each with gene expression profiles over time. We observed a temporal uncoupling: antibody correlates peaked at day 1, linked to innate and antiviral pathways,^41,54^ and did not overlap with those linked to T cell responses. In contrast, T cell correlates were weaker and peaked at day 3, with intriguing pre-vaccination signatures. Benchmarking against mRNA vaccines showed that robust innate induction is necessary but insufficient for strong humoral immunity; deeper regulatory and metabolic programs appear essential for optimal antibody responses.

The diverse range of antibody responses across our cohorts—from minimal induction with inhaled MVA, to variable intramuscular MVA, to strong mRNA vaccine responses—provided a broad, continuous spectrum. This enabled our machine learning approach to leverage the full granularity of both transcriptional and serological data, mapping individual gene expression patterns to antibody outcomes. This high-resolution molecular mapping of vaccine immunogenicity captured subtle regulatory differences, revealing targetable molecular pathways for vaccine optimization. For instance, CD274 (PD-L1), an immune checkpoint molecule and established clinical target,^58^ was associated with strong antibody responses in our study, while GAS6 highlights TAM/Axl signaling as a potentially modulatory axis for fine-tuning vaccine-induced immunity.^59^

Despite implementation challenges, inhaled vaccines offer a promising alternative to overcome limitations of parenteral vaccines.^55^ Our study provides a molecular characterization of inhaled vaccination in humans, demonstrating that inhaled delivery engages immune programs fundamentally similar to intramuscular vaccination, while simultaneously inducing local T cell immunity with reduced systemic inflammation. This compartmentalized response provides a compelling molecular rationale for mucosal vaccine strategies. The temporal uncoupling between transcriptional correlates of humoral and cellular responses, alongside identification of predictive genes, refines our understanding of vaccine immunogenicity determinants and offers actionable insights for future vaccine design.

We acknowledge several limitations. The small sample size inherent to phase 1 trials limits statistical power and generalizability, and only a single dose level of the inhaled vaccine was evaluated due to safety considerations. Multi-omics profiling was not performed uniformly across inhaled and intramuscular cohorts, restricting direct comparisons in some modalities. Finally, the controlled inhalation protocol^33^ used here is not scalable to large vaccination efforts, though the broader feasibility of inhaled vaccines has been demonstrated in multiple phase 3 and 4 trials.^15,22,60,61^

In conclusion, our in-depth analysis of human immune responses to inhaled vaccination provides a molecular foundation and immediate targets for data-driven mucosal vaccine development, highlighting the value of systems approaches in guiding rational vaccine design.

## Methods

### Study design and participants

To investigate immune responses induced by mucosal vaccination against SARS-CoV-2, we conducted a clinical trial assessing an inhaled MVA-ST vaccine in previously immunized adults. For comparative analysis of transcriptional responses following systemic versus mucosal vaccine delivery, we additionally analyzed blood transcriptomes from a parallel clinical trial in which the same vaccine was administered via intramuscular injection.

#### Inhaled vaccine cohort

This phase 1 trial (NCT05226390) was conducted at Hannover Medical School to evaluate the safety, reactogenicity, and immunogenicity of MVA-ST administered via inhalation. The study was approved by the National Competent Authority (Paul Ehrlich Institute; EudraCT number: 2020-004010-35) and the Ethics Committee at Hannover Medical School (EK 10012_AMG_mono_2021). Written informed consent was obtained from all participants. Eligible individuals had previously received three doses of any EU-approved SARS-CoV-2 vaccine (last dose ≥3 months prior to enrollment). Participants (n=23) received a single inhaled dose of 10^7^ IU MVA-ST in 0.5 mL. The vaccine is based on a MVA vector encoding a prefusion-stabilized SARS-CoV-2 Spike protein.^25^ Full protocol details, including eligibility criteria, inhalation delivery procedures, participant characteristics, safety, and immunological outcomes have been reported elsewhere.^33,34^

#### Intramuscular comparator cohort

To compare transcriptional responses following intramuscular vaccination, we analyzed samples from participants in Part B of a separate phase 1b trial (NCT04895449) conducted at the University Medical Center Hamburg-Eppendorf. The study was approved by the Paul Ehrlich Institute (EudraCT number: 2021-00548-23) and the Ethics Committee of the Hamburg Medical Association (2021-100621-AMG-ff), and written informed consent was obtained from all participants. In Part B of the trial, individuals with two prior doses of BNT162b2 (last dose ≥6 months prior to enrollment) received a single intramuscular injection of MVA-ST at one of three dose levels: 10^7^ IU (nC=C12), 5C×C10^7^C±C0.5Clog IU (nC=C10), or 10^8^C±C0.5Clog IU (nC=C8). Detailed methods, participant characteristics, and immunological outcomes have been described previously.^32^

### Sample Collection and Processing

Peripheral blood and BAL samples were collected from participants at defined time points relative to vaccination, as shown in Fig. 1b. Blood was drawn into EDTA and heparin tubes; plasma was separated by centrifugation and stored at –80°C for cytokine measurements. Peripheral blood mononuclear cells (PBMCs) were isolated via density gradient centrifugation and analyzed fresh for downstream applications. BAL samples were collected by bronchoscopy using standard lavage procedures, filtered, and centrifuged to separate cells and supernatant. BAL cells were analyzed fresh, while BAL fluid aliquots were stored at – 80°C for cytokine profiling.

### Spectral Flow Cytometry

Fresh whole blood samples were collected in EDTA K3E S-Monovettes (Sarstedt) and processed in duplicate. One mL of blood was diluted into 20 mL red blood cell lysis buffer (168.25 mmol NH4Cl, 10 mmol KHCO3, 1mmol EDTA-Na2 in 1l ddH2O; pH7.3) and incubated for 20 min at room temperature. After two PBS washes, cells were stained for 20 min at room temperature using two antibody panels (Supplementary Tables 7–8), washed again, and acquired on a Cytek Aurora spectral flow cytometer (5-laser configuration: 355, 405, 488, 561, 640 nm). Data were recorded using SpectroFlow (Cytek, version 3.0.3). Dead cells and cell doublets were excluded using FCS Express 7 (De Novo Software). For downstream analysis, cells were pre-gated as CD45+CD3-CD19-(innate panel), and CD45+CD3+ or CD45+CD19+ (adaptive panel), and exported as .fcs files.

Unsupervised high-dimensional analysis was performed using a previously established R pipeline.^62,63^ In brief, pre-gated .fcs files were imported^64^, asinh-transformed^65^, and batch-corrected^66^. Cells were assigned to cell clusters using FlowSOM^67^ based on canonical marker expressions. To reduce redundancy while preserving biological resolution, highly similar clusters—differing only by minor shifts in non-informative markers—were merged after manual inspection.^68,69^ Contaminant populations were excluded, and remaining clusters were manually annotated based on global marker characteristics. Dimensionality reduction was performed with UMAP, and the relative abundance of each cluster per sample was calculated for statistical analysis.

### Cytokine Profiling

Plasma and BAL fluid samples were prepared for multiplex cytokine analysis as previously described.^62^ Cytokine measurements were performed using the Bio-Plex Pro Human Cytokine Screening Panel (48-Plex, #12007283, Bio-Rad, USA) according to the manufacturer’s protocol. Plates were read on a Bio-Plex 200 System (#171000201, Bio-Rad, USA). To minimize technical variability, all samples were processed in parallel on the same day and measured the following day. For plasma, only cytokines with ≥80% of values falling within the quantifiable range of the standard curve were retained to ensure robust statistical analysis of reliably quantifiable analytes. Values below the lower limit of detection were set to half the minimum detectable concentration. In BAL fluid, where the majority of analytes fell below detection limits, all analytes were retained to allow exploratory profiling given the limited availability of multiplex cytokine data in this compartment.

### Interferon-γ release assay (IGRA)

S1-specific T cell responses were assessed using an interferon-γ release assay (Euroimmun, #ET 2606-3003). Lithium-heparinized whole blood was stimulated for 20-24h, after which plasma interferon-γ concentrations were quantified by ELISA (Euroimmun. #EQ 6841-9601) following the manufacturer’s instructions. Optical density values were acquired on a SpectraMax iD3 plate reader (Molecular Devices, San Jose, USA) and analyzed using SoftMax Pro v7.1.2.W

### PBMC and BAL Cell Stimulation

PBMCs and BAL cells were restimulated with a SARS-CoV-2 peptide pool as previously described.^70^ For intracellular cytokine staining, cells were incubated in complete RPMI medium supplemented with brefeldin A (Sigma-Aldrich) to inhibit cytokine secretion and allow intracellular accumulation. In parallel, for multiplex cytokine measurements of secreted analytes, cells were stimulated under identical conditions but without brefeldin A. DMSO at equivalent volumes was used as a negative control in all assays. Cytokine values from peptide-stimulated samples were background-corrected by subtracting corresponding DMSO control values. Background-corrected values below zero were set to the lower limit of detection for the respective analyte to avoid biologically implausible negative concentrations. Only analytes showing a clear response above background—CXCL9, CXCL10, and IFN-γ— were subjected to formal statistical testing.

### Whole blood RNA Sequencing and Analysis

#### Sample collection, RNA extraction, sequencing

Whole blood was collected from study participants in PAXgene RNA tubes (QIAGEN) and processed according to manufacturer’s instructions. Tubes were gently inverted immediately after collection and held at room temperature for approximately 2 hours before storage at –80LJ°C. Total RNA was extracted from thawed samples using the PAXgene Blood RNA Kit (QIAGEN) following the manufacturer’s protocol, and stored at –80LJ°C until use. RNA sequencing libraries were generated using the Illumina TruSeq mRNA protocol. Sequencing was performed on an Illumina NovaSeq X Plus platform (2LJ×LJ61LJbp) and NovaSeq 6000 platform (2LJ×LJ100LJbp) for the Hannover and Hamburg cohorts, respectively.

#### Data processing and analysis

Transcript quantification was performed using kallisto^71^ (v0.51) with pseudo-alignment to the human reference transcriptome (Ensembl Release 114). Genes with low expression were filtered using the *filterByExpr* function from the edgeR package.^72^ Batch correction across datasets was performed using ComBat-Seq,^73^ and its effectiveness was evaluated by principal component analysis (PCA) before and after correction. Differential gene expression analysis was conducted using the limma-voom workflow, accounting for repeated measures within individuals via the *duplicateCorrelation* function.^74^ P-values were adjusted for multiple testing using the Benjamini-Hochberg method. Blood transcription module (BTM)^41^ enrichment analysis was performed using the CERNO test, as implemented in the tmod package.^75^ Module activity was assessed using module eigengenes, defined as the first principal component of gene expression within each module, capturing the dominant expression pattern for all genes in that module.^42^ Transcription factor activity was inferred using the decoupleR package^76^, leveraging prior regulatory networks to estimate transcription factor enrichment scores from the gene expression matrix based on the CollecTRI database. Concordance and discordance scores were calculated based on gene log_2_-fold changes and adjusted *P* values obtained from prior differential expression analyses, as previously described.^77^

#### Integration of external mRNA vaccine cohorts

To contextualize the transcriptional responses in our cohort, we analyzed published RNA-seq datasets from independent cohorts who had received mRNA-based SARS-CoV-2 vaccines. Details for each cohort—including vaccination type (prime/boost), sampling schedule, number of participants and samples, GEO accession numbers, and references—are summarized in Supplementary Table 3.

### Machine Learning and Predictive Modeling

We employed a two-step machine learning framework to identify early transcriptional predictors of subsequent antibody responses, defined as the log_2_-fold change of S1-specific IgG titers at day 28 relative to day 0. In the first step, gene-level log_2_-fold changes between day 0 and day 1 were filtered by variance, retaining the top 50%, and used to train an initial XGBoost^43^ model with 5-fold cross-validation over 20 random splits (15 boosting rounds). This lightweight configuration was chosen to reduce computational complexity and limit overfitting, as the primary goal of this stage was feature preselection. By narrowing the high-dimensional input space, we aimed to limit noise and enhance the efficiency of downstream modeling. Features were ranked by their mean SHapley Additive exPlanations (SHAP) values^44^ across folds, and the top 1000 genes were retained. In the second step, we trained a more computationally intensive model on the reduced feature space using 10-fold cross-validation repeated over 50 iterations (50 boosting rounds). This final model was designed for outcome prediction; the use of more folds and iterations was motivated by the small sample size, allowing more efficient data use while reducing variance in performance estimation. Model performance in the training cohort was assessed using out-of-fold (OOF) predictions. The trained model was also evaluated on data not used during training: a held-out subset of 8 randomly selected samples from the MVA cohort, and two independent second-dose mRNA vaccine cohorts (mRNA 2nd (A) and mRNA 2nd (HH; see Supplementary Table 3)). Second-dose mRNA vaccine datasets were used because all available third-dose mRNA datasets had been included in the training to maximize model development. Within the mRNA 2nd (HH) cohort, only individuals not overlapping with the mRNA 3^rd^ (HH) cohort were included to ensure full independence of the test dataset. Model performance was evaluated using Pearson correlations between predicted and observed values, as well as root mean squared error (RMSE) and mean absolute error (MAE). Feature importance in the final predictive model was also interpreted using SHAP values, allowing identification of the most influential genes contributing to antibody response prediction.

### Statistical Analysis

All statistical analyses were conducted using R (version 4.4) and Python (version 3.10).^78,79^ Two-sided *P* values <0.05 were considered statistically significant. Where appropriate, *P* values were adjusted for multiple testing using the Benjamini-Hochberg false discovery rate (FDR) method. Unpaired group comparisons were performed using the Wilcoxon rank-sum test; for comparisons involving more than two groups, the Kruskal-Wallis test was applied, followed by Dunn’s post hoc test with FDR correction. Paired comparisons were analyzed using the Wilcoxon signed-rank test for two-group comparisons and the Friedman test for comparisons involving more than two related measurements, followed by pairwise Wilcoxon signed-rank tests with FDR adjustment, where applicable. Differential testing of log-transformed cytokine concentrations and immune cell cluster frequencies across study time points was performed using linear modeling with empirical Bayes moderation, as implemented in the *limma* framework.^74^ Paired comparisons were modeled using a design matrix accounting for repeated measures within participants. Correlation analyses were performed using Spearman’s rank correlation unless stated otherwise. Data visualizations were created using established frameworks within the R environment.^80,81^. Full details on statistical tests used are provided in the figure legends.

## Data availability

RNA-seq data from the Hannover and Hamburg trials have been deposited in the Gene Expression Omnibus (GEO) repository under the accession numbers GSE291673 and GSE291862, respectively. Accession numbers for external RNA-seq datasets are provided in Supplementary Table 6. Other data are available from the corresponding authors upon reasonable request.

## Code availability

All analyses were performed using standard, publicly available software and packages as detailed in the Methods. Custom scripts used for data processing and analysis are available from the corresponding author upon reasonable request.

## Acknowledgements

This work was supported by funds of the State of Lower Saxony (14-76103-184 CORONA-11/20), the German Center for Infection Research (Deutsches Zentrum für Infektionsforschung, DZIF (TTU 01.934, TTU 01.941, TTU 01.924, TTU 01.709), the German Research Foundation (Deutsche Forschungsgemeinschaft, DFG) Excellence Strategy EXC 2155 “RESIST” (Project ID39087428), the German Center for Lung Research (Deutsches Lungenzentrum, DZL, grant 82DZL002B1), and the DFG Research Infrastructure NGS_CC (project 407495230) as part of the Next Generation Sequencing Competence Network (project 423957469). NGS analyses of the Hannover trial samples were performed at the Research Core Unit Genomics (RCUG) at Hannover Medical School, and NGS analyses of the Hamburg trial samples at the Competence Centre for Genomic Analysis Kiel (CCGA). We thank Thomas Hesterkamp, Anna Kutschenko, Christoph Schindler, Mark Permanyer, Anika Buchholz and Antonia Zapf for their contributions to the study, and Monika Friedrich for preparing and coordinating the RNA sequencing of the Hamburg samples with the CCGA Kiel. L.R. was supported by the Hannover Biomedical Research School (HBRS) and the Center for Infection Biology (ZIB), the Joachim Herz Foundation, the German Center for Lung Research, reand the Else Kröner-Fresenius-Stiftung (Fördervertrag 2018_Kolleg.12). Fig. 1a and 6a were created in BioRender.

## Author contributions

Conceptualization (clinical trials): J.H., M.M.A., A.B., A.Z.; Conceptualization (systems analysis): L.R., I.O., S.H., R.F.; Methodology: A.V., G.S., V.K., S.B., J.H., M.M.A., L.R., I.O., S.H., R.F.; Investigation: S.H., R.G., I.O., J.B.M., M.F., I.R., J.S., A.R., L.M.W., L.M., V.K., S.B., R.K., C.F., A.V.; Data curation and formal analysis: L.R., A.H., I.O., R.F., Visualization: L.R.; Funding acquisition: R.F., J.H., M.M.A.; Resources: R.F., G.H., S.B., C.F., J.H., M.M.A.; Supervision: R.F.; Writing (original draft): L.R., R.F.; Writing (review & editing): All authors reviewed and approved the final manuscript.

