## Supplementary figures and images for "Systems Vaccinology Reveals Distinct Immune Signatures of Inhaled and Intramuscular SARS-CoV-2 Vaccination in Humans"

### Suppl. Fig 1

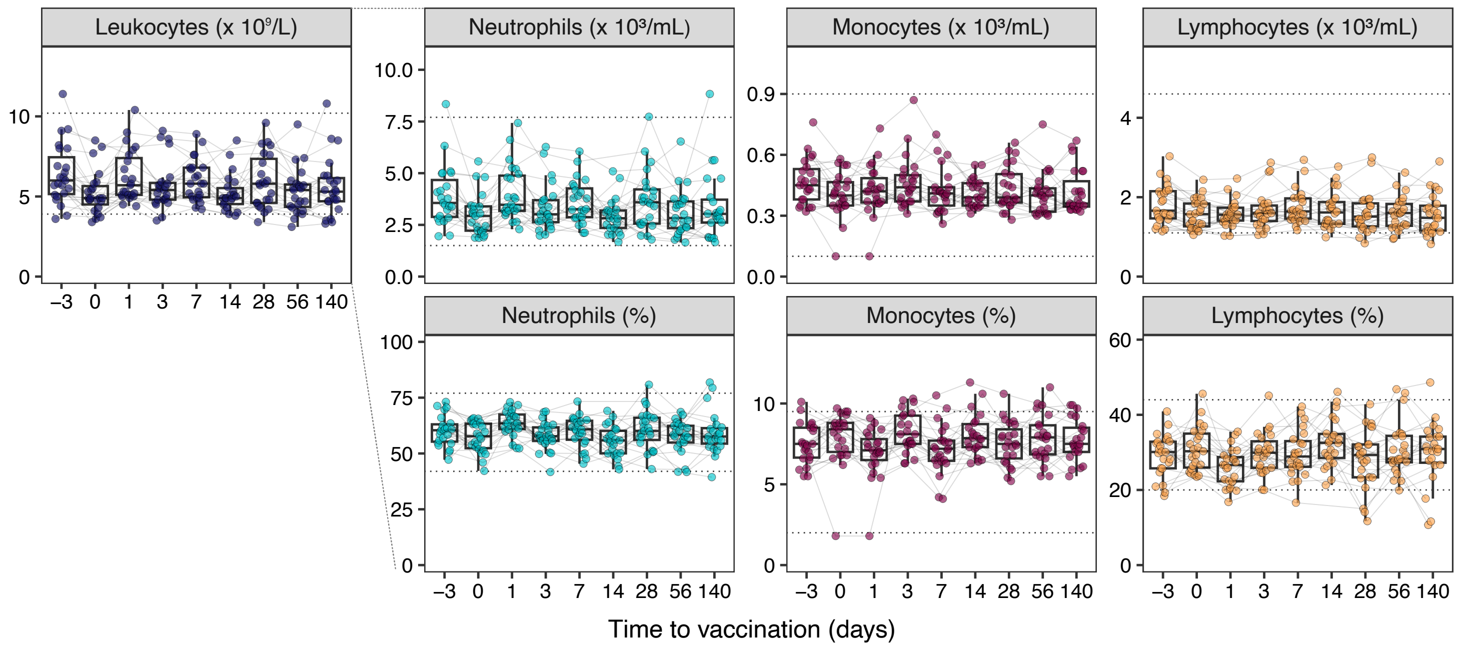

### Suppl. Fig 2

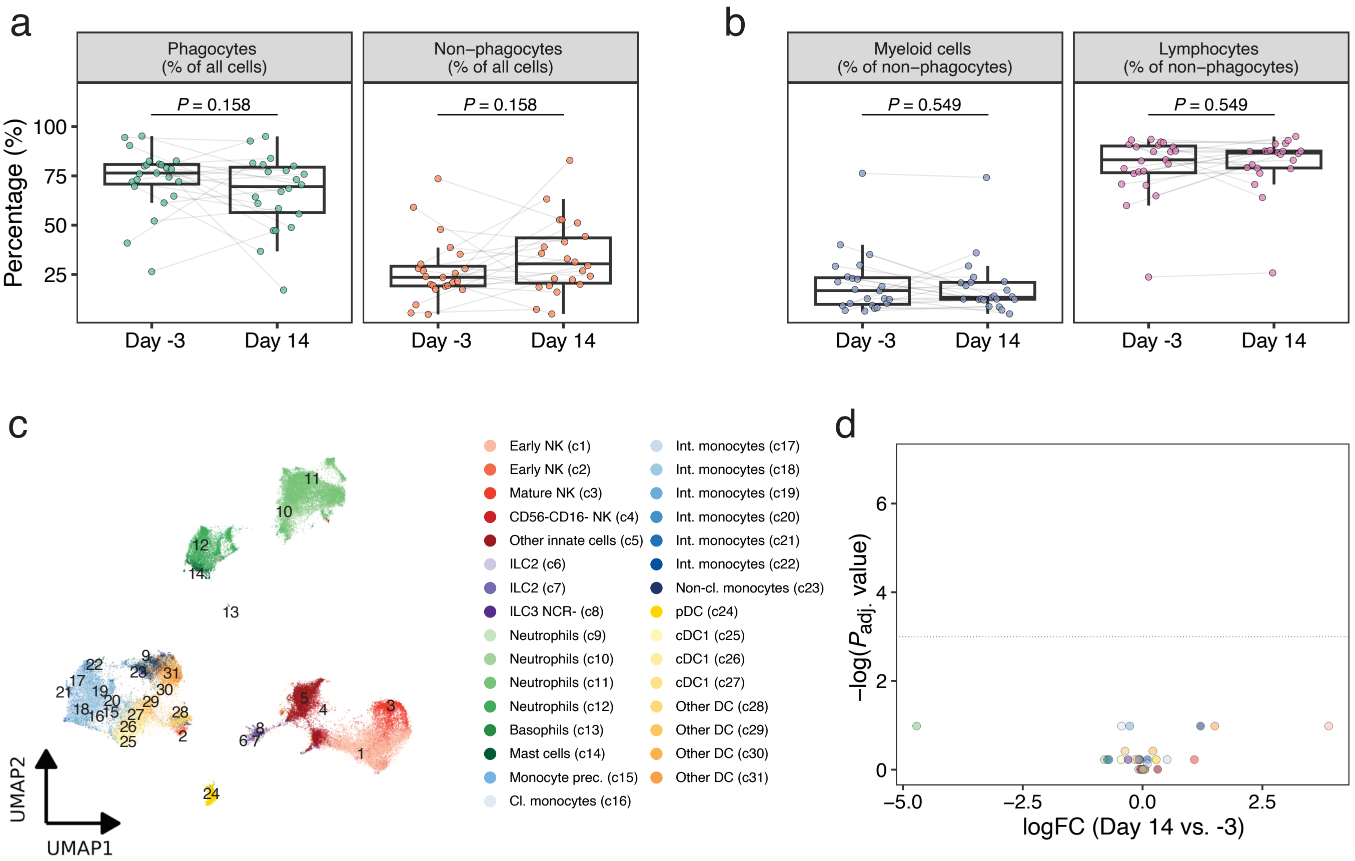

### Suppl. Fig 3

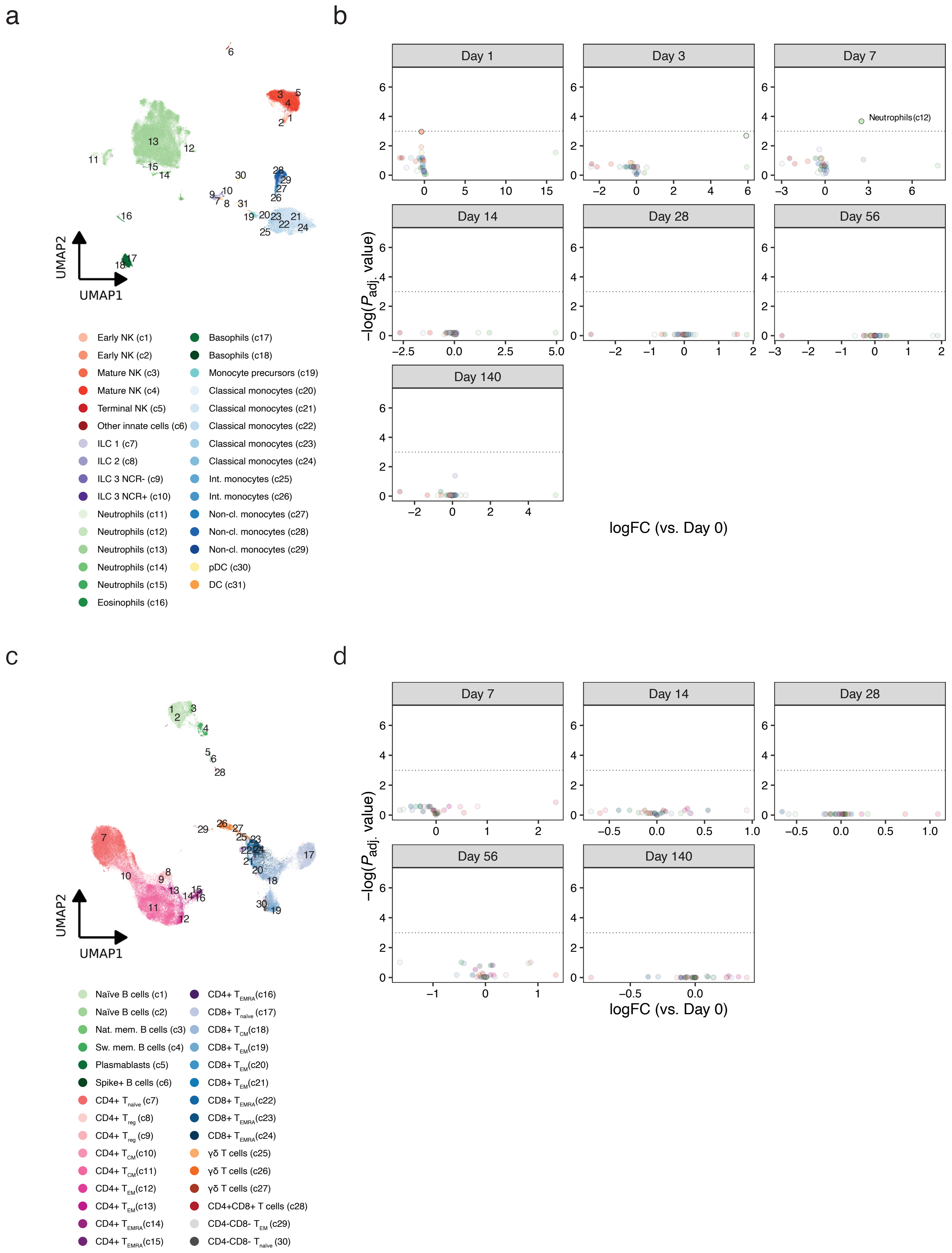

### Suppl. Fig 4

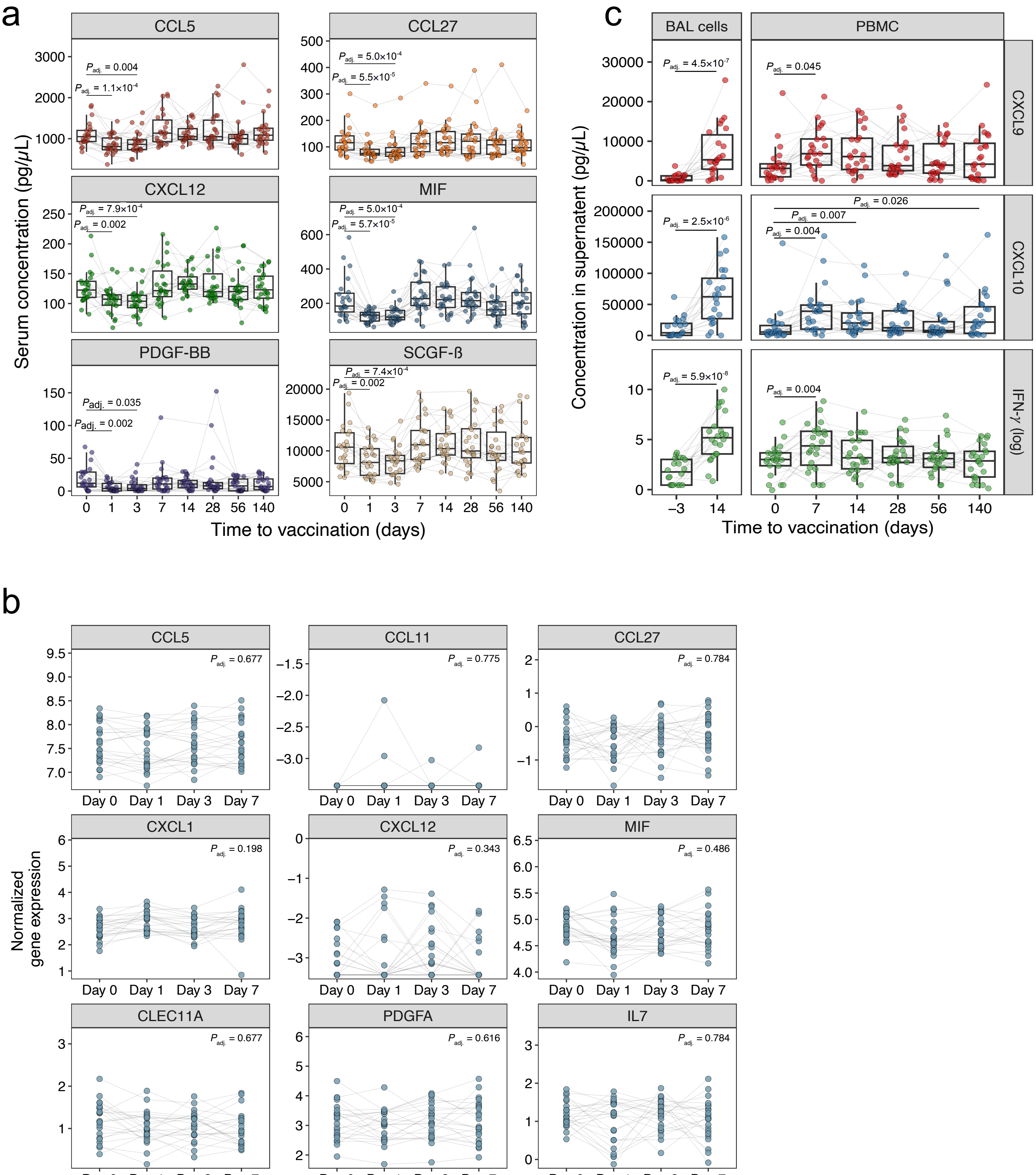

### Suppl. Fig 5

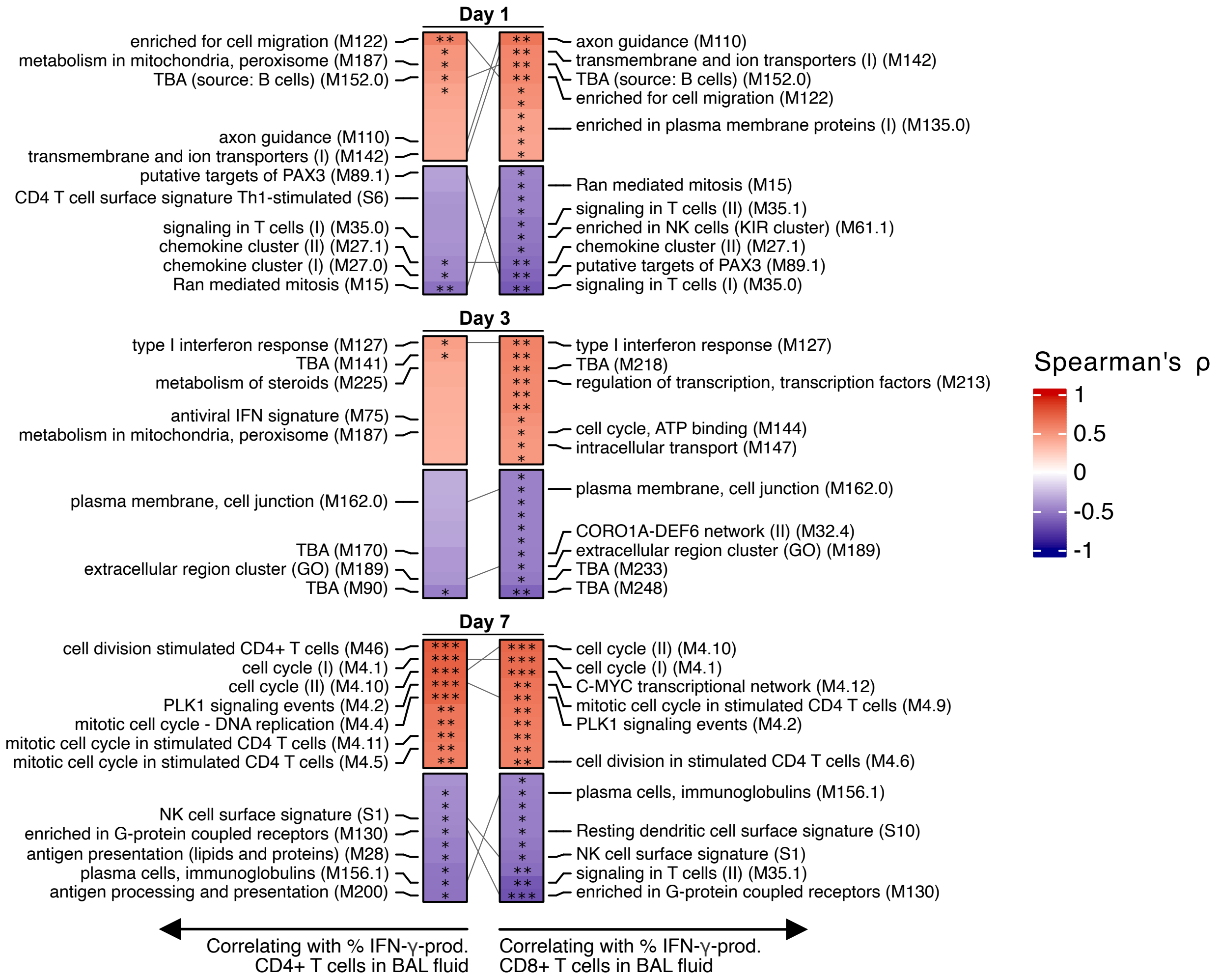

### Suppl. Fig 6

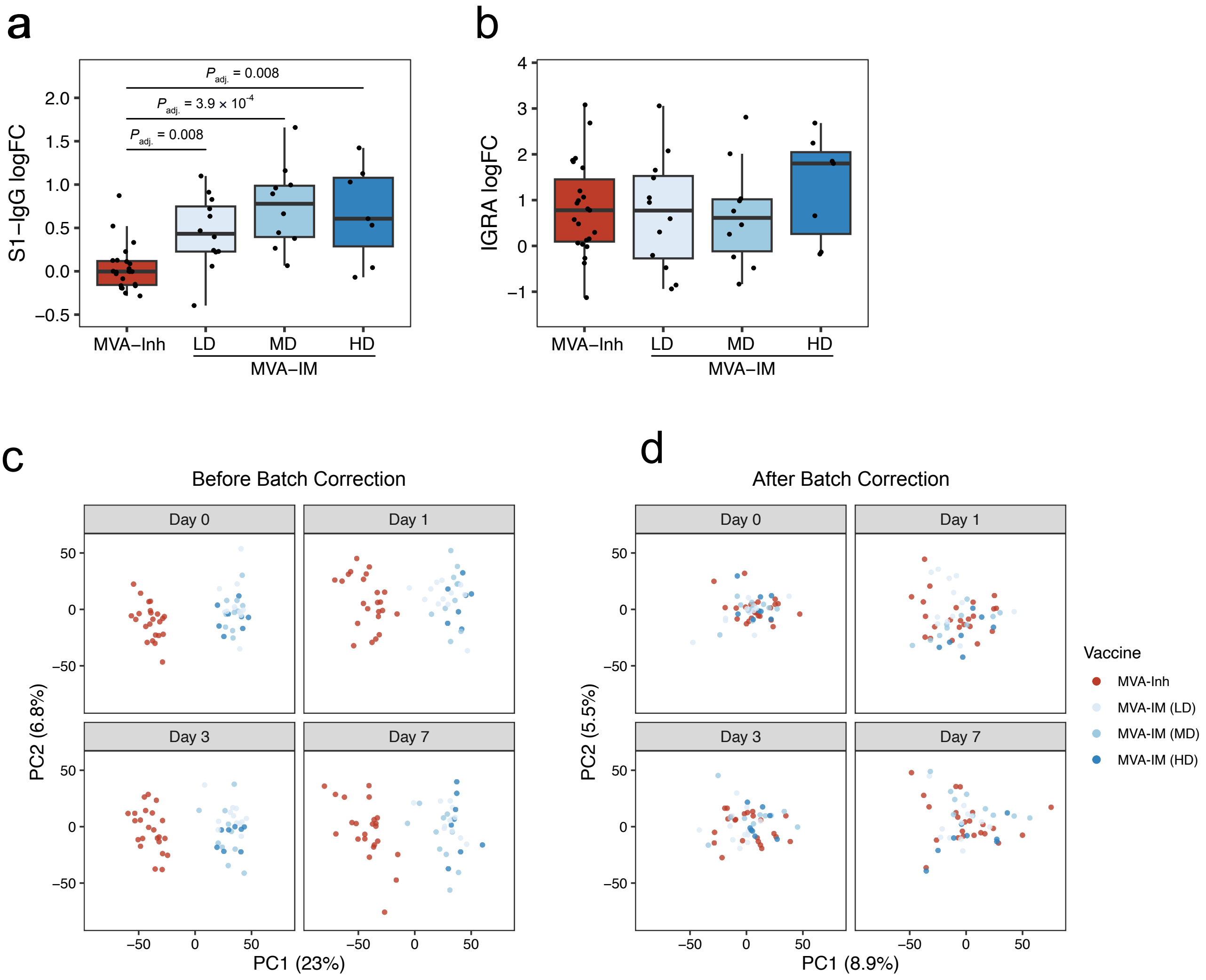

### Suppl. Fig 7

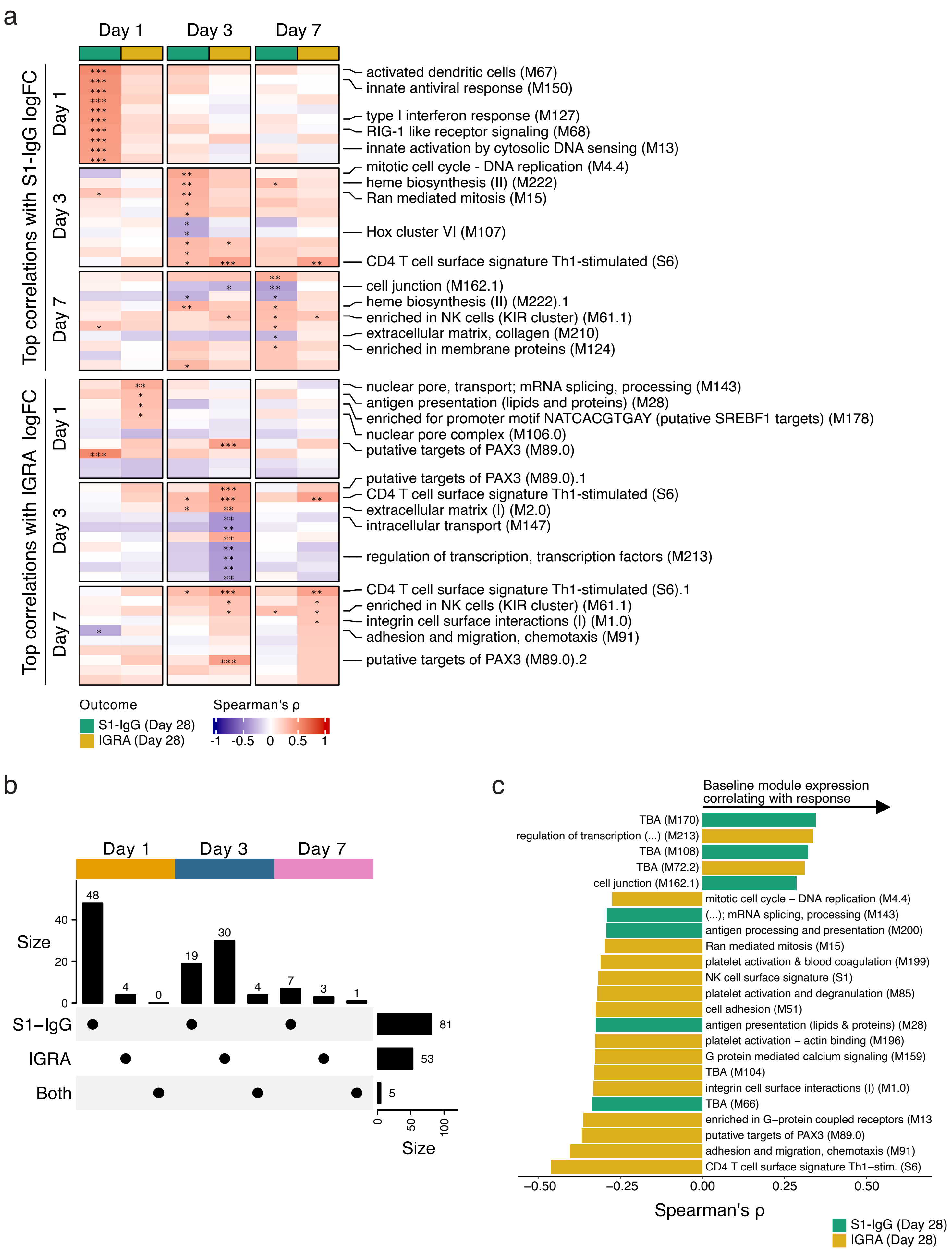

### Suppl. Fig 8

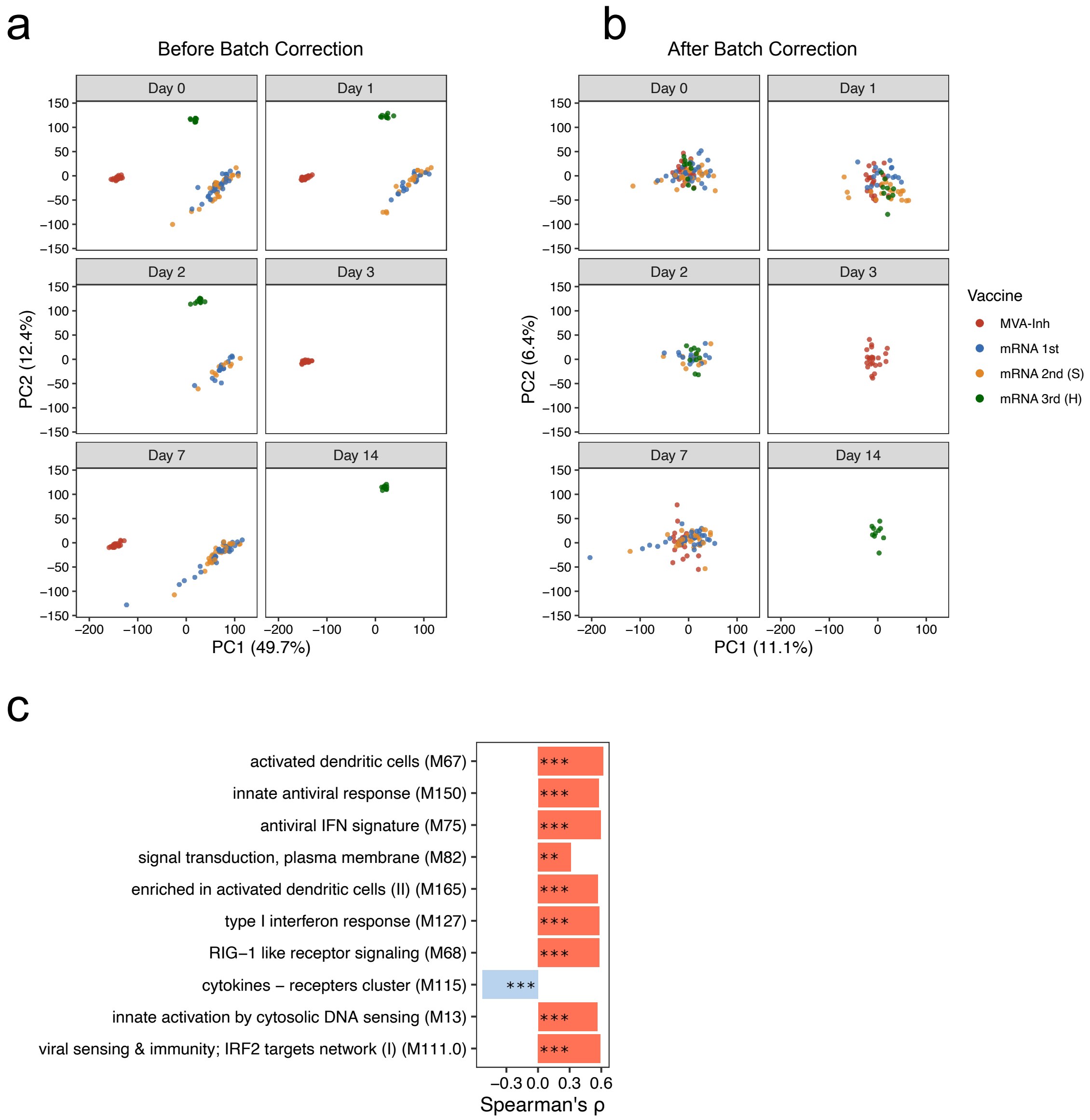

### Suppl. Fig 9

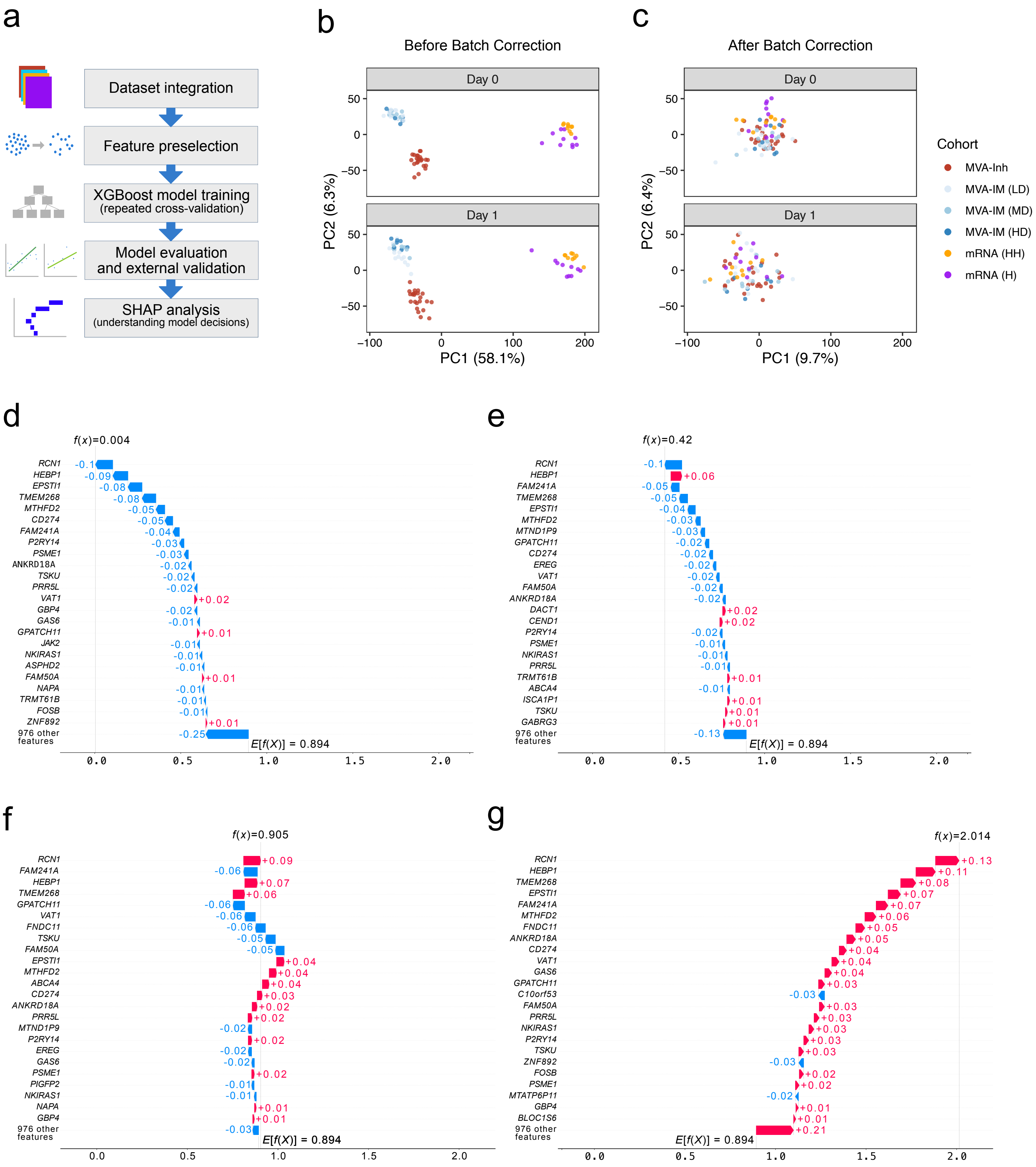
