## Supplement Fig Legends for "Systems Vaccinology Reveals Distinct Immune Signatures of Inhaled and Intramuscular SARS-CoV-2 Vaccination in Humans"

**Supplementary Figure Legends**

**Supplementary Fig. 1: Clinical hematology parameters following MVA vaccination.** Hematological safety monitoring. Longitudinal changes in peripheral blood leukocyte subsets, including total leukocyte counts and absolute/relative frequencies of neutrophils, monocytes, and lymphocytes. Dashed lines indicate clinical reference ranges. Box plots represent medians and interquartile ranges at each time point.

**Supplementary Fig. 2: Immune cell changes in bronchoalveolar lavage fluid after inhaled vaccination.**

**a,** Frequencies of phagocytic (left) and non-phagocytic (right) cells as a proportion of total BAL cells at baseline (day -3) and post-vaccination (day 14), shown as box plots with individual data points. **b**, Proportions of myeloid and lymphoid cells within the non-phagocytic BAL compartment at the same time points. Statistical comparisons in **a** and **b** were performed using paired Wilcoxon tests. **c**, UMAP projection of 31 innate immune clusters identified by unsupervised clustering of non-phagocytic BAL cells. **d**, Differential abundance analysis of innate immune clusters in non-phagocytic BAL cells, shown as a volcano plot. The dashed line indicates an adjusted *P* value threshold of 0.05.

**Supplementary Fig. 3: Immune cell changes in peripheral blood after inhaled vaccination. a**, UMAP projection of 31 innate immune cell clusters identified by unsupervised clustering of fresh peripheral blood samples. **b**, Differential abundance analysis of innate immune cell clusters at post-vaccination time points compared to baseline, shown as volcano plots. The dashed line indicates an adjusted *P* value threshold of 0.05. **c**, UMAP projection of 30 adaptive immune cell clusters identified by unsupervised clustering of fresh peripheral blood samples. **d**, Differential abundance analysis of adaptive immune cell clusters at post-vaccination time points compared to baseline, shown as volcano plots. The dashed line indicates an adjusted *P* value threshold of 0.05.

**Supplementary Fig. 4: Cytokine- and chemokine-related gene-expression dynamics.**

**a**, Full longitudinal course of all available time points for cytokines and chemokines that were significantly altered at days 1 and 3, showing their temporal dynamics from baseline through day 140. **b**, Normalized gene expression of genes encoding cytokines and chemokines that were found to be significantly altered at the protein level on days 1 and 3 post-vaccination. No significant longitudinal differences were observed across time points. Statistical analysis was performed using the Friedman test. **c**, CXCL9, CXCL10, and IFN-γ concentrations in culture supernatants from BAL cells and PBMCs after *ex vivo* restimulation with a SARS-CoV-2 Spike peptide pool. BAL samples were collected only before (day -3) and after (day 14) vaccination, whereas PBMCs were also collected at later time points

**Supplementary Fig. 5: Correlation of blood transcriptional modules with IFN-γ-producing T cells in the lung.**

Spearman correlations between changes in blood transcriptional modules on days 1 (top), 3 (middle), and 7 (bottom) post-vaccination and the frequency of IFN-γ-producing CD4+ (left columns) and CD8+ (right columns) T cells in BAL fluid sampled at day 14. For each time point, the top 10 most significant positive (upper) and negative (lower) correlations are displayed. Lines connecting heatmap columns indicate shared gene modules between CD4+ and CD8+ T cell analyses. Key modules are labelled for clarity. * *P* <0.05, ** *P* < 0.01, *** *P* < 0.001.

**Supplementary Fig. 6: Comparison of transcriptional responses between inhaled and intramuscular MVA vaccine delivery.**

**a**, S1-specific IgG responses in the inhaled MVA group and three intramuscular MVA groups (low, medium, and high dose). Box plots show log_2_-fold changes in antibody titers between day 0 to day 28. **b**, IFN-γ responses measured by IFN-γ release assay (IGRA) in the same groups, shown as show log_2_-fold changes between day 0 to day 28. Differences across groups in **a** and **b** were assessed using the Kruskal-Wallis test followed by Dunn’s post hoc test with Benjamini–Hochberg adjustment for multiple comparisons. **c**, Principal component analysis (PCA) of whole-blood gene expression across the four groups prior to batch correction. **d**, PCA after batch correction, demonstrating effective mitigation of batch effects.

**Supplementary Fig. 7: Transcriptional correlates of antibody and T cell responses.**

**a**, Heatmap showing Spearman correlation coefficients between changes in blood transcriptional module eigengene expression (at days 1, 3, and 7 post-vaccination) and downstream immunological outcomes: log_2_-fold change in S1-specific IgG titers (day 28 vs. 0) and IFN-γ release from peptide-stimulated PBMC cultures (day 28 vs. 0). Per time point, the top 10 most significant modules associated with each time point are shown. The upper half of each heatmap displays modules ranked by correlation with antibody responses, and the lower half by correlation with cellular responses; in both cases, corresponding correlations with the alternate outcome are also shown. Asterisks indicate statistical significance (* *P* <0.05, ** *P* < 0.01, *** *P* < 0.001). Inhaled and IM vaccine groups were jointly analyzed based on their concordant transcriptional responses. **b**, Upset plot showing the number of blood transcriptional modules whose expression changes from baseline at days 1, 3, and 7 significantly correlated with antibody (S1-IgG) responses, T cell (IGRA) responses, or both. The highest number of significant correlations was observed at day 1 for antibody responses. **c**, Spearman correlation coefficients between pre-vaccination blood transcriptional module expression and antibody (green) and cellular (orange) responses. Only significant correlations are shown.

**Supplementary Fig. 8: Transcriptomic integration and validation across inhaled MVA and mRNA vaccines.**

**a**, Principal component analysis (PCA) of whole-blood transcriptomic profiles from participants receiving inhaled MVA vaccine and mRNA vaccines (first, second, or third dose), prior to batch correction. **b**, PCA after batch correction, showing effective mitigation of batch effects. **c**, Validation of correlation patterns from Fig. 6 in an expanded dataset including mRNA vaccine recipients. Nine of the ten top blood transcriptional modules associated with antibody responses at day 1 retained the same direction and statistical significance; one module reversed direction.

**Supplementary Fig. 9: Predictive modeling of antibody responses across vaccine cohorts. a**, Schematic overview of the machine learning workflow used to predict S1-specific antibody responses at day 28 based on gene-wise log_2_-fold changes from day 0 to day 1 post-vaccination. **b**, Principal component analysis (PCA) of transcriptomic data from included cohorts in this analysis prior to batch correction. Datasets used for independent validation were excluded from the batch correction step to avoid data leakage. **c**, PCA after batch correction, demonstrating effective mitigation of batch effects across cohorts. **d–g**, SHAP waterfall plots showing ensemble-averaged feature contributions for representative individual participants from **d**, the inhaled MVA cohort; **e**, low-dose MVA-IM cohort (same dose as inhaled); **f**, medium-dose MVA-IM cohort; and **g**, mRNA vaccine cohort (third dose). Each bar represents the average SHAP value across all 500 XGBoost models (50 iterations × 10 folds), indicating how each feature (gene) contributes to shifting the model’s prediction above or below the ensemble baseline, E[f(X)] (i.e., the average predicted log_2_-fold change in S1-specific IgG across all cohorts included in this analysis). The final prediction, f(x), corresponds to the sum of the baseline plus all feature contributions, representing the predicted antibody response for each participant. This approach enables identification of features that most strongly influence predicted antibody responses, providing participant-level interpretability. The observed antibody log_2_-fold changes for the selected participants were 0.00, 0.40, 0.89, and 2.01 for panels **d–g**, respectively. Individuals were chosen to be representative of group-level means in each cohort.
