## Supplement Tables for "Systems Vaccinology Reveals Distinct Immune Signatures of Inhaled and Intramuscular SARS-CoV-2 Vaccination in Humans"

**Supplementary Table 1: Innate immune cell clusters (BAL fluid).**

| **Cluster** | **Subset** | **Marker characteristics** |
| --- | --- | --- |
| 1 | Early NK | CD16-CD56^high^CD94+ |
| 2 | Early NK | CD16-CD56^high^CXCR3+CD94^low^CD33^low^ |
| 3 | Mature NK | CD16+CD56+CD94+NKG2D^high^ |
| 4 | CD56-CD16- NK | CD16-CD56-NKG2D^high^ |
| 5 | Other innate cells | Lin-CD16-CD127+NKp46+ |
| 6 | ILC2 | Lin-CD127+CD117+CRTH2^high^NKG2D+CD115+ |
| 7 | ILC2 | Lin-CD127+CD117+CRTH2+NKG2D^high^CD115+ |
| 8 | ILC3 NCR- | Lin-CD127+CD117+NKG2D^low^NKp46+CXCR3^high^ |
| 9 | Neutrophils | CD16+CD66b+HLA-DR^pos^ |
| 10 | Neutrophils | CD16^high^CD66b+CXCR3+HLA-DR^neg^ |
| 11 | Neutrophils | CD16^high^CD66b^high^CXCR3+HLA-DR^neg^ |
| 12 | Eosinophils | CD16-CD66b+CRTH2+CCR3+ |
| 13 | Basophils | CD123^high^CCR3^high^FCeRIa^high^CRTH2^high^ |
| 14 | Mast cells | CD33+CD123^low^FCeRIa^low^ |
| 15 | Monocyte precursors | CD16-CD14^low^CD33^high^CCR2-CD11c+HLA-DR^high^CD86^high^ |
| 16 | Classical monocytes | CD16-CD14^high^CD33^high^CCR2+CD11c+HLA-DR^pos^ |
| 17 | Int. monocytes | CD16+CD14^high^CD33^high^CCR2+CD11c+HLA-DR^pos^CD86^high^CD206+ |
| 18 | Int. monocytes | CD16+CD14+CD33+CD11c+HLA-DR^pos^CD206+ |
| 19 | Int. monocytes | CD16+CD14^high^CD33^high^CCR2^high^CD11c+HLA-DR^pos^ |
| 20 | Int. monocytes | CD16+CD14+CD33^high^CD206+FCeRIa+ |
| 21 | Int. monocytes | CD16+CD14+CD33^high^HLA-DR^pos^CD11c^high^CRTH2+CXCR3+ |
| 22 | Int. monocytes | CD16+CD14^high^CD33^high^HLA-DR^pos^CD11c^high^CRTH2+CXCR3- |
| 23 | Non-cl. monocytes | CD16+CD14-CD33+HLA-DR^pos^ |
| 24 | pDC | Lin-CD16-CD123^high^CXCR3+HLA-DR^pos^ |
| 25 | cDC1 | Lin-CD16-FCeRIa^high^CD11c^high^CD206+ |
| 26 | cDC2 | Lin-CD16-FCeRIa^high^CD11c^high^CD33+CD206+ |
| 27 | cDC2 | Lin-CD16^low^FCeRIa^high^CD11c^high^CD33+CD206+ |
| 28 | Other DC | Lin-CD16-FCeRIa-CD11c^high^CXCR3+ |
| 29 | Other DC | Lin-CD16-FCeRIa-CD11c^high^CD86^high^ |
| 30 | Other DC | Lin-CD16-HLA-DR^high^ |
| 31 | Other DC | Lin-CD16^low^HLA-DR^high^ |

**Supplementary Table 2: Adaptive immune cell clusters (BAL fluid)**

| **Cluster** | **Subset** | **Marker characteristics** |
| --- | --- | --- |
| 1 | Trans. B cells | CD19+CD20+IgG-IgM+IgD^low^CD27-CD10^low^Spike+ |
| 2 | Trans. B cells | CD19+CD20+CD16^high^IgG^high^IgM+IgD^low^CD27-CD10^high^Spike+ |
| 3 | Trans. B cells | CD19+CD20+IgG+IgM+IgD+CD27+CCR7+CD28+CD10+  CXCR5^high^Spike^high^ |
| 4 | Nat. mem. B cells | CD19^high^CD20^high^CD45RA+IgG-IgM-IgD^high^CD27+CCR7+CD21^high^CD24^high^CXCR5^high^ |
| 5 | CD4+ T_naive_ | CD3+CD4+CD62L+CCR7+CD45RA+CD27+ |
| 6 | CD4+ T_reg_ | CD3+CD4+CD27-CD28+CD45RA-CD25+CD95+ |
| 7 | CD4+ T_reg_ | CD3+CD4+CD27+CD28+CD45RA-CCR7+CD25+CD95+ |
| 8 | CD4+ T_CM_ | CD3+CD4+CD27+CD28+CD45RA-CCR7+CD95+ |
| 9 | CD4+ T_CM_ | CD3+CD4+CD27-CD45RA-CCR7+CD56+CXCR3+CTLA4+ |
| 10 | CD4+ T_CM_ | CD3+CD4+CD27+CD28+CD45RA-CCR7+ |
| 11 | CD4+ T_EM_ | CD3+CD4+CD27-CD45RA- |
| 12 | CD4+ T_EM_ | CD3+CD4+CD27-CD28+CD45RA- |
| 13 | CD4+ T_EM_ | CD3+CD4+CD27-CD28-CD45RA- |
| 14 | CD4+ T_naive_ | CD3+CD8+CD27+CD28+CD45RA+CCR7+ |
| 15 | CD8+ T_cytotoxic_ | CD3+CD8+CD27-CD28-CD45RA-CD56^high^ |
| 16 | CD8+ T_cytotoxic_ | CD3+CD8+CD27+CD28+CD45RA-CD56^high^ |
| 17 | CD8+ T_cytotoxic_ | CD3+CD8+CD27+CD28^low^CD45RA-CD56+ |
| 18 | CD8+ T_EM_ | CD3+CD8+CD27-CD28-CD45RA- |
| 19 | CD8+ T_EM_ | CD3+CD8+CD27+CD28^low^CD45RA- |
| 20 | CD8+ T_EM_ | CD3+CD8+CD27-CD28-CD45RA- |
| 21 | CD8+ T_EMRA_ | CD3+CD8+CD27+CD28^low^CD45RA+ |
| 22 | CD8+ T_EMRA_ | CD3+CD8+CD27-CD28-CD45RA+ |
| 23 | γδ T cells | CD3+TCRgd+CD4+CD27+CD28+CD45RA-CCR7+CCR4^high^CXCR5+CTLA4+Spike^high^ |
| 24 | γδ T cells | CD3+TCRgd+CD27+CD45RA+ |
| 25 | γδ T cells | CD3+TCRgd+CD8+CD27+CD45RA-CCR7+CCR4+CTLA4+Spike+ |
| 26 | γδ T cells | CD3+TCRgd+CD8+CD27+CD45RA-CCR7+CCR4+CD56+CXCR3+CTLA4+Spike+ |
| 27 | γδ T cells | CD3+TCRgd+CD27-CD45RA+CD56^high^ |

**Supplementary Table 3: Innate immune cell clusters (blood).**

| **Cluster** | **Subset** | **Marker characteristics** |
| --- | --- | --- |
| 1 | Early NK | CD16^low^CD56^high^CD94^high^NKp46+NKG2D^high^CD127+ |
| 2 | Early NK | CD16-CD56^high^CD94^high^NKp46+NKG2D^high^CD127+ |
| 3 | Mature NK | CD16+CD56+CD94^low^ |
| 4 | Mature NK | CD16+CD56+CD94^low^NKG2D+ |
| 5 | Terminal NK | CD16+CD56-CD94^low^NKG2D+ |
| 6 | Other innate cells | CD16+CD56-CD94^high^NKp46^high^CD127+CD115+  CD86+CD80+CD66b+ |
| 7 | ILC 1 | Lin-CD16-CD127+CD117^low^ |
| 8 | ILC 2 | Lin-CD16-CD127+CD117+CRTH2^low^ |
| 9 | ILC 3 NCR- | Lin-CD16^low^CD127+CD117+ |
| 10 | ILC 3 NCR+ | Lin-CD16-CD127+CD117+NKG2D+ |
| 11 | Neutrophils | CD66b+CD125+CD16^high^+CD11c+CD80+ |
| 12 | Neutrophils | CD66b+CD125+CD16^high^+CD11c+ |
| 13 | Neutrophils | CD66b+CD125+CD16^high^+CD11c+HLA-DR^low^ |
| 14 | Neutrophils | CD66b+CD16^high^+CD80+ |
| 15 | Neutrophils | CD66b^high^CD16- |
| 16 | Eosinophils | CD66b+CD117+CD16-CCR3+CRTH2+ |
| 17 | Basophils | Lin-CD123^high^CCR3^high^FCeRIa^high^CRTH2^high^ |
| 18 | Basophils | Lin-CD123^high^CCR3^high^FCeRIa+CRTH2^high^ |
| 19 | Monocyte precursors | CD14-CD16-CD33+HLA-DR^high^FCeRIa+CD11c+ |
| 20 | Cl. monocytes | CD14+CD16-CD33+HLA-DR^pos^FCeRIa+CD11c+CD163^high^ |
| 21 | Cl. monocytes | CD14+CD16-CD33+HLA-DR^pos^CD11c+ |
| 22 | Cl. monocytes | CD14+CD16-CD33+HLA-DR^pos^CD11c+ |
| 23 | Cl. monocytes | CD14^high^CD16-CD33+HLA-DR^high^CD163^high^ |
| 24 | Cl. monocytes | CD14+CD16-CD33+HLA-DR^low^ |
| 25 | Int. monocytes | CD14+CD16+CD33+CD125^high^ |
| 26 | Int. monocytes | CD14+CD16+CD33+CD125+HLA-DR^high^ |
| 27 | Int. monocytes | CD14^low^CD16+CD33+CD125+HLA-DR^high^ |
| 28 | Non-cl. monocytes | CD14-CD16+CD33+CD11c^high^CD86^high^ |
| 29 | Non-cl. monocytes | CD14-CD16+CD33+CD11c^high^CD86^high^ |
| 30 | pDC | Lin-CXCR3+CD123^high^CCR2+HLA-DR^higha^ |
| 31 | DC | Lin-HLA-DR^high^CD117+CD11c+ |

**Supplementary Table 4: Adaptive immune cell clusters (blood).**

| **Cluster** | **Subset** | **Marker characteristics** |
| --- | --- | --- |
| 1 | Naïve B cells | CD19+CD20+IgG-IgM+IgD+CD27- |
| 2 | Naïve B cells | CD19+CD20+IgG+IgM+IgD+CD27- |
| 3 | Nat. mem. B cells | CD19+CD20+IgG-IgM+IgD+CD27+ |
| 4 | Sw. mem. B cells | CD19+CD20+IgG+IgM-IgD-CD27+ |
| 5 | Plasmablasts | CD19+CD20-IgG-IgM-IgD-CD27+ |
| 6 | Spike-spec. B cells | CD19+CD20+IgG^high^IgM+IgD-CD27-CD16^high^CTLA4+Spike^high^ |
| 7 | CD4+ T_naive_ | CD3+CD4+CD45RA+CCR7+CD62L+CD27+CD28+ |
| 8 | CD4+ T_reg_ | CD3+CD4+CD25+CD45RA-CCR7+CD62L+CD27+CD28+CCR4+CD127+ |
| 9 | CD4+ T_reg_ | CD3+CD4+CD25+CD45RA-CCR7+CD62L+CD27+CD28+CCR4+ |
| 10 | CD4+ T_CM_ | CD3+CD4+CD45RA+CCR7+CD62L+  CD27+CD28+CD45RO+ |
| 11 | CD4+ T_CM_ | CD3+CD4+CD45RA-CCR7+CD62L+CD27+CD28+ |
| 12 | CD4+ T_EM_ | CD3+CD4+CD45RA-CCR6+CD27-CD28+ |
| 13 | CD4+ T_EM_ | CD3+CD4+CD45RA-CXCR3+CD27+CD28+ |
| 14 | CD4+ T_EMRA_ | CD3+CD4+CD45RA+CD27-CD28-CD45RO+ |
| 15 | CD4+ T_EMRA_ | CD3+CD4+CD45RA+CD27-CD28+CD45RO+ |
| 16 | CD4+ T_EMRA_ | CD3+CD4+CD45RA+CD27-CD28-CD56^high^CD45RO+ |
| 17 | CD8+ T_naive_ | CD3+CD8+CD45RA+CCR7+CD62L+CD27+CD28+ |
| 18 | CD8+ T_CM_ | CD3+CD8+CD45RA-CCR7^low^CD27+CD28+ |
| 19 | CD8+ T_EM_ | CD3+CD8+CD45RA-CCR6+CD27+CD28+CD56+ |
| 20 | CD8+ T_EM_ | CD3+CD8+CD45RA-CD27-CD28- |
| 21 | CD8+ T_EM_ | CD3+CD8+CD45RA-CD27-CD28-CD56+ |
| 22 | CD8+ T_EMRA_ | CD3+CD8+CD45RA+CD27-CD28-CD56+ |
| 23 | CD8+ T_EMRA_ | CD3+CD8+CD45RA+CD27-CD28- |
| 24 | CD8+ TEMRA | CD3+CD8+CD45RA+CD27^low^ |
| 25 | γδ T cells | CD3+TCRgd+CD45RA+CD27-CD28-CD56+ |
| 26 | γδ T cells | CD3+TCRgd+CD45RA-CD27+CD28+ |
| 27 | γδ T cells | CD3+TCRgd+CD45RA+CD27-CD28-CD56+CD127- |
| 28 | CD4+CD8+ T cells | CD3+CD4+CD8+CD45RA+CD27+CD28+CD45RO+CCR4+CXCR5+CTLA4+Spike+ |
| 29 | CD4+CD8+ T_EM_ | CD3+CD45RA-CD27+CD28+CD56+CCR6+ |
| 30 | CD4+CD8+ T_naive_ | CD3+CD45RA+CD27+CD28+CCR7+ |

**Supplementary Table 5: Multiplex cytokine profiling (BAL fluid).**

| Analyte | Proportion <OOR (%) | Concentration at day -3 (pre- vaccination) | Concentration at day 14 (post-vaccination) | *P* value | *P*_adj._ value |
| --- | --- | --- | --- | --- | --- |
| CCL2 | 64.4 | 0.1 (0.1-0.3) | 0.1 (0.1-0.5) | 0.799 | 1.000 |
| CCL3 | 73.3 | 0 (0-0) | 0 (0-0.2) | 0.050 | 0.597 |
| CCL4 | 86.7 | 0 (0-0) | 0 (0-0) | 0.855 | 1.000 |
| CCL5 | 31.1 | 10.1 (2.9-25.5) | 8.9 (0.3-14.1) | 0.097 | 0.597 |
| CCL7 | 100.0 | 0.6 (0.6-0.6) | 0.6 (0.6-0.6) | - | - |
| CCL11 | 95.6 | 0 (0-0) | 0 (0-0) | 1.000 | 1.000 |
| CCL27 | 95.6 | 0 (0-0) | 0 (0-0) | 1.000 | 1.000 |
| CXCL1 | 11.1 | 344.8 (204-590.6) | 765 (272.8-835.6) | 0.102 | 0.597 |
| CXCL8 | 0.0 | 7.7 (4.7-13.4) | 13.4 (7.3-20) | 0.079 | 0.597 |
| CXCL9 | 73.3 | 0.2 (0.2-0.3) | 0.2 (0.2-0.3) | 0.594 | 1.000 |
| CXCL10 | 6.7 | 24.4 (9.6-37.4) | 28.6 (20.3-72.8) | 0.022 | 0.597 |
| CXCL12 | 100.0 | 7.6 (7.6-7.6) | 7.6 (7.6-7.6) | - | - |
| IFN-α2 | 100.0 | 0.1 (0.1-0.1) | 0.1 (0.1-0.1) | - | - |
| IFN-**γ** | 71.1 | 0 (0-0) | 0 (0-0.3) | 0.359 | 1.000 |
| IL-1a | 93.3 | 0.1 (0.1-0.1) | 0.1 (0.1-0.1) | 1.000 | 1.000 |
| IL-1b | 91.1 | 0 (0-0) | 0 (0-0) | 0.586 | 1.000 |
| IL-1RA | 2.2 | 80 (54.1-126.3) | 85.7 (56.2-153.1) | 0.224 | 0.979 |
| IL-2 | 100.0 | 0 (0-0) | 0 (0-0) | - | - |
| IL-2Rα | 97.8 | 0 (0-0) | 0 (0-0) | 1.000 | 1.000 |
| IL-3 | 100.0 | 0 (0-0) | 0 (0-0) | - | - |
| IL-4 | 71.1 | 0 (0-0) | 0 (0-0) | 0.782 | 1.000 |
| IL-5 | 100.0 | 5.4 (5.4-5.4) | 5.4 (5.4-5.4) | - | - |
| IL-6 | 57.8 | 0 (0-0.2) | 0.1 (0-0.3) | 0.286 | 1.000 |
| IL-7 | 97.8 | 0.3 (0.3-0.3) | 0.3 (0.3-0.3) | 1.000 | 1.000 |
| IL-9 | 100.0 | 1.3 (1.3-1.3) | 1.3 (1.3-1.3) | - | - |
| IL-10 | 97.8 | 0 (0-0) | 0 (0-0) | 1.000 | 1.000 |
| IL-12(p40) | 100.0 | 0.3 (0.3-0.3) | 0.3 (0.3-0.3) | - | - |
| IL-12(p70) | 82.2 | 0 (0-0) | 0 (0-0) | 1.000 | 1.000 |
| IL-13 | 97.8 | 0 (0-0) | 0 (0-0) | 1.000 | 1.000 |
| IL-15 | 100.0 | 3.6 (3.6-3.6) | 3.6 (3.6-3.6) | - | - |
| IL-16 | 15.6 | 3.3 (0.3-6.6) | 5.2 (2.9-9.5) | 0.178 | 0.888 |
| IL-17 | 95.6 | 0 (0-0) | 0 (0-0) | 1.000 | 1.000 |
| IL-18 | 11.1 | 2.4 (0.9-5.4) | 1.8 (1-3.4) | 0.808 | 1.000 |
| FGF-β | 97.8 | 0.3 (0.3-0.3) | 0.3 (0.3-0.3) | 1.000 | 1.000 |
| GCSF | 0.0 | 13.5 (9.2-22.4) | 15.7 (10-29.9) | 0.074 | 0.597 |
| GMCSF | 97.8 | 0 (0-0) | 0 (0-0) | 1.000 | 1.000 |
| HGF | 100.0 | 0.4 (0.4-0.4) | 0.4 (0.4-0.4) | - | - |
| LIF | 97.8 | 0.2 (0.2-0.2) | 0.2 (0.2-0.2) | 1.000 | 1.000 |
| MCSF | 88.9 | 0 (0-0) | 0 (0-0) | 1.000 | 1.000 |
| MIF | 0.0 | 579.4 (325.6-821.3) | 534.7 (325.8-928.3) | 0.679 | 1.000 |
| PDGF-bb | 100.0 | 0 (0-0) | 0 (0-0) | - | - |
| SCF | 55.6 | 0 (0-0.7) | 0 (0-0.7) | 0.505 | 1.000 |
| SCGF-β | 40.0 | 181.8 (41.3-602.6) | 338.7 (41.3-515.4) | 0.809 | 1.000 |
| β-NGF | 82.2 | 0 (0-0) | 0 (0-0) | 0.573 | 1.000 |
| TNF-α | 95.6 | 0.4 (0.4-0.4) | 0.4 (0.4-0.4) | 1.000 | 1.000 |
| TNF-β | 100.0 | 0 (0-0) | 0 (0-0) | - | - |
| TRAIL | 53.3 | 0.1 (0.1-0.5) | 0.2 (0.1-0.8) | 0.551 | 1.000 |
| VEGF | 100.0 | 5 (5-5) | 5 (5-5) | - | - |

**Supplementary Table 6: Datasets used for cross-comparison with mRNA vaccination.**

| **Cohort name** | **Vaccine** | **Vaccination** | **Time points (days)** | **Participants (n)** | **Samples (n total)** | **Reference and GEO accession** |
| --- | --- | --- | --- | --- | --- | --- |
| MVA-Inh | Inhaled MVA vaccine | Third  (2nd boost) | 0,1,3,7 | N=23 | N=92 | This study  (GSE291673) |
| MVA-IM | Intramuscular MVA vaccine  (LD, MD, HD) | Third  (2nd boost) | 0,1,3,7 | N=30 | N=119 | This study  (GSE291862) |
| mRNA 1st | mRNA vaccine (BNT162b) | Prime | 0,1,2,7 | N=32  (Day 1: 16,  Day 2: 15) | N=94 | Arunachalam et al.^1^  (GSE169159) |
| mRNA 2nd (S) | mRNA vaccine (BNT162b) | Second  (1st boost) | 0,1,2,7 | N = 31  (Day 1: 20,  Day 2: 10) | N=91 | Arunachalam et al.^1^  (GSE169159) |
| mRNA 2nd (HH) | mRNA vaccine (BNT162b; 2x mRNA-1273) | Second  (1st boost) | 0,1 | N = 7 | N=14 | Odak et al.^2^  (GSE247401) |
| mRNA 3rd (H) | mRNA vaccine (BNT162b) | Third  (2nd boost) | 0,1,3,14 | N=10 | N=38 | Odak et al.^2^  (GSE246525) |
| mRNA 3rd (HH) | mRNA vaccine (BNT162b; 2x mRNA-1273) | Third  (2nd boost) | 0,1 | N=10 | N=20 | Odak et al.^2^  (GSE247401) |

S = Stanford, HH = Hamburg, H = Hannover (recruitment sites)

1	Arunachalam PS, Scott MKD, Hagan T, *et al.* Systems vaccinology of the BNT162b2 mRNA vaccine in humans. *Nature* 2021; 596: 410–6.

2	Odak I, Riemann L, Sandrock I, *et al.* Systems biology analysis reveals distinct molecular signatures associated with immune responsiveness to the BNT162b COVID-19 vaccine. *EBioMedicine* 2024; 99. DOI:10.1016/j.ebiom.2023.104947.

**Supplementary Table 7: Innate immune cell antibody panel.**

| **Specificity** | **Fluorochrome** | **Clone** | **Company** | **Cat#** | **Dilution** |
| --- | --- | --- | --- | --- | --- |
| **CD45** | BUV395 | HI30 | BD | 563792 | 1:200 |
| **CD16** | BUV496 | 3G8 | BD | 612944 | 1:100 |
| **CXCR3** | BUV563 | 1C6/CXCR3 | BD | 741406 | 1:50 |
| **CD115 (CSF-1R)** | BUV615 | 9-4D2-1E4 | BD | 751279 | 1:50 |
| **CD11c** | BUV661 | B-ly6 | BD | 612967 | 1:100 |
| **CD56** | BUV737 | NCAM16.2 | BD | 612766 | 1:100 |
| **CD86** | BUV805 | BU63 | BD | 748375 | 1:50 |
| **CD123** | Super Bright 436 | 6H6 | Invitrogen | 62-1239-42 | 1:50 |
| **CD94** | BV480 | HP-3D9 | BD | 746737 | 1:20 |
| **CD33** | BV510 | WM53 | BioLegend | 303422 | 1:20 |
| **HLA-DR** | BV570 | L243 | BioLegend | 307638 | 1:50 |
| **CCR3** | BV605 | 5E8 | BioLegend | 310716 | 1:20 |
| **CD80** | BV650 | 2D10 | BioLegend | 305227 | 1:100 |
| **CD163** | BV711 | GHI/61 | BioLegend | 333630 | 1:50 |
| **CD125** | BV786 | A14 | BD | 743932 | 1:100 |
| **FCeRIa** | FITC | AER-37 (CRA-1) | BioLegend | 334608 | 1:100 |
| **CD3** | AF532 | UCHT1 | Invitrogen | 58-0038-42 | 1:50 |
| **CD14** | BB700 | MφP9 | BD | 566465 | 1:200 |
| **NKp46** | PerCP Cy5.5 | 9E2 | BioLegend | 331920 | 1:20 |
| **NKg2D (CD314)** | PE | 1D11 | BioLegend | 320805 | 1:100 |
| **CRTH2 (CD294)** | PE-Dazzle 594 | BM16 | BioLegend | 350125 | 1:50 |
| **CD117** | PE Cy5 | A3C6E2 | BioLegend | 323412 | 1:20 |
| **CD19** | PE-Cy7 | HIB19 | BioLegend | 982410 | 1:200 |
| **CCR2 (CD192)** | APC | K036C2 | BioLegend | 357208 | 1:100 |
| **CD66b** | AF647 | G10F5 | Biolegend | 305110 | 1:100 |
| **CD127** | AF700 | A019D5 | BioLegend | 351344 | 1:100 |
| **viability** | Zombie NIR |  | Biolegend | 423106 | 1:400 |
| **CD206** | APC/Fire750 | 15-2 | BioLegend | 321134 | 1:50 |

**Supplementary Table 8: Adaptive immune cell antibody panel.**

| **Specificity** | **Fluorochrome** | **Clone** | **Company** | **Cat#** | **Dilution** |
| --- | --- | --- | --- | --- | --- |
| **CD45RA** | BUV395 | HI100 | BD | 740298 | 1:200 |
| **CD16** | BUV496 | 3G8 | BD | 612944 | 1:100 |
| **CD4** | BUV563 | RPA-T4 | BD | 741353 | 1:200 |
| **CCR4** | BUV615 | 1G1 | BD | 613000 | 1:20 |
| **CD21** | BUV661 | 1048 | BD | 750187 | 1:100 |
| **CD56** | BUV737 | NCAM16.2 | BD | 612766 | 1:100 |
| **CD27** | BUV805 | L128 | BD | 569167 | 1:100 |
| **CD28** | BV421 | CD28.2 | Biolegend | 302930 | 1:100 |
| **CD45RO** | Pacific Blue | UCHL1 | Biolegend | 304216 | 1:100 |
| **IgD** | BV480 | IA6-2 | BD | 566138 | 1:200 |
| **CD19** | BV510 | HIB19 | Biolegend | 302242 | 1:200 |
| **IgM** | BV570 | MHM-88 | Biolegend | 314518 | 1:200 |
| **CD62L** | BV605 | DREG-56 | Biolegend | 304834 | 1:200 |
| **CXCR3** | BV650 | G025H7 | Biolegend | 353730 | 1:100 |
| **CCR6** | BV711 | G034E3 | Biolegend | 353436 | 1:100 |
| **CXCR5** | BV750 | RF8B2 | BD | 747111 | 1:200 |
| **CCR7** | BV785 | G043H7 | Biolegend | 353230 | 1:50 |
| **Spike** | mNEONgreen | Produced by T. Krey | | | 5 µg/sample |
| **CD20** | AF532 | 2H7 | Invitrogen | 58-0209-42 | 1:200 |
| **CD8** | SparkBlue550 | SK1 | Biolegend | 344760 | 1:200 |
| **CD14** | BB700 | MφP9 | BD | 566465 | 1:100 |
| **CD38** | PerCP-eF710 | HB7 | Invitrogen | 46-0388-42 | 1:100 |
| **CD10** | PE | HI10a | Biolegend | 312204 | 1:100 |
| **CD24** | PE/Dazzle 594 | ML5 | Biolegend | 311134 | 1:100 |
| **CD95** | PE-Cy5 | DX2 | Biolegend | 305610 | 1:20 |
| **CD25** | PE-Fire700 | M-A251 | Biolegend | 356146 | 1:100 |
| **CTLA4** | PE-Cy7 | L3D10 | Biolegend | 349914 | 1:20 |
| **HLA-DR** | PE-Fire810 | L243 | Biolegend | 307683 | 1:50 |
| **TCRgd** | APC | 11F2 | Miltenyi | 130-113-500 | 1:50 |
| **IgG Fc** | AF647 | M1310G05 | Biolegend | 410714 | 1:200 |
| **CD127** | APC-R700 | A019D5 | Biolegend | 351344 | 1:100 |
| **viability** | Zombie NIR |  | Biolegend | 423106 | 1:400 |
| **Tim3** | APC Fire750 | F38-2E2 | Biolegend | 345044 | 1:100 |
| **CD3** | APC Fire810 | SK7 | Biolegend | 344858 | 1:50 |
